# Systematic Review Association between Endotype of Prematurity and Mortality: A Systematic Review, Meta-analysis and Meta-regression

**DOI:** 10.1101/2023.01.21.23284854

**Authors:** Tamara M Hundscheid, Eduardo Villamor-Martinez, Eduardo Villamor

**Affiliations:** Department of Pediatrics, Maastricht University Medical Center (MUMC+), School for Oncology and Reproduction (GROW), Maastricht, the Netherlands; Statistics Netherlands, 6412HX Heerlen, the Netherlands

**Author notes:** Corresponding Author: Eduardo Villamor, Department of Pediatrics, Maastricht University Medical Center (MUMC+), Postbus 5800, 6202 AZ Maastricht, the Netherlands.

**Keywords:** preterm birth, chorioamnionitis, hypertensive disorders of pregnancy, growth restriction, mortality

## Abstract

**Introduction:** Preterm birth represents the leading cause of neonatal mortality. Pathophysiological pathways, or endotypes, leading to prematurity can be clustered into infection/inflammation and dysfunctional placentation. We aimed to perform a systematic review and meta-analysis exploring the association between these endotypes and risk of mortality during first hospital admission.

**Methods:** PROSPERO ID: CRD42020184843. PubMed and Embase were searched for observational studies examining infants with gestational age (GA) ≤34 weeks. Chorioamnionitis represented the infectious-inflammatory endotype, while dysfunctional placentation proxies were hypertensive disorders of pregnancy (HDP) and small for GA (SGA)/intrauterine growth restriction (IUGR). A random-effects model was used to calculate odds ratios (ORs) and 95% confidence intervals (CIs). Heterogeneity was studied using random-effects meta-regression analysis.

**Results:** Of 4322 potentially relevant studies, 150 (612,580 infants) were included. Meta-analysis showed a positive mortality odds for chorioamnionitis (OR 1.43, 95% CI 1.25-1.62) and SGA/IUGR (OR 1.68, 95% CI 1.38-2.04), but a negative mortality odds for HDP (OR 0.74, 95% CI 0.64-0.86). Chorioamnionitis was associated with a lower GA, while HDP and SGA/IUGR were associated with a higher GA. Meta-regression showed a significant correlation between these differences in GA and mortality odds.

**Discussion/Conclusion:** Our data suggest that the infectious/inflammatory endotype of prematurity has a greater overall impact on mortality risk as it is the most frequent endotype in the lower GAs. However, when the endotype of placental dysfunction is severe enough to induce growth restriction, it is strongly associated with higher mortality rates even though newborns are more mature.

## Introduction

Preterm birth is defined by the World Health Organization as delivery before 37 completed weeks of gestational age (GA) and is further subdivided into extreme (less than 28 weeks), very (28 to less than 32 weeks) moderate (32 to less than 34 weeks), and late (34 to less than 37 weeks) preterm birth. Although survival has increased substantially in the last few decades, preterm birth and its associated complications represent the leading cause of both neonatal and childhood mortality worldwide (1-3).

Although GA is the main prognostic factor in prematurity, there is growing recognition that the pathological conditions triggering preterm birth play a central role in the development of its complications (4-8). McElrath et al. proposed two main pathways to extreme preterm birth: 1) infection/inflammation and 2) dysfunctional placentation. The first group include chorioamnionitis, pre-labor premature rupture of membranes, placental abruption, and cervical insufficiency (9). The second group is characterized by the presence of histologic features of placental vascular dysfunction and is associated with hypertensive disorders of pregnancy (HDP) and the entity identified as fetal indication/intrauterine growth restriction (IUGR) (9). These two etiological pathways possess the characteristics to be considered as the main endotypes of very and extreme preterm birth (4, 7).

The term endotype refers to ‘‘a subtype of a condition, which is defined by a distinct functional or pathophysiological mechanism.’’(10). Endotypes are thus a different form of classification from clinical phenotypes and describe distinct disease entities with a defining etiology and/or a consistent pathophysiological mechanism. In line with the approach of personalized medicine, endotyping will facilitate the design of more targeted therapeutic and prognostic approaches (4, 11-14).

Despite the growing awareness about the role of the pathophysiologic mechanism, or endotype, triggering prematurity in its outcome (4-7, 15-19), this variable is rarely taken into account in the models predicting mortality in the preterm newborn (20-22). Furthermore, to the best of our knowledge, the association between the endotype of prematurity and mortality during primary hospital admission has not been systematically reviewed. The purpose of the present study was to address this gap in the literature. Following the methodology described in our previous meta-analyses (7, 23), the infectious-inflammatory endotype was represented by chorioamnionitis, while the dysfunctional placentation endotype was represented by HDP and IUGR. Since many researchers use small for gestational age (SGA) as a proxy for IUGR, and although the two conditions are not necessarily synonymous (24-27), we also collected data on SGA. From now on, this group will be referred to as SGA/IUGR. In addition, through the use of meta-regression, we aimed to unravel the role of variables such as GA, infant sex, or use of antenatal corticosteroids in the association between endotype of prematurity and mortality.

## Methods

The study was conducted according to the PRISMA and MOOSE guidelines. Detailed information on methods is provided as supplementary material. Review protocol was registered in PROSPERO database (ID= CRD42020184843). The Population, Exposure, Comparison and Outcome (PECO) question was: Do preterm infants (P) exposed to chorioamnionitis, HDP, or growth restriction during pregnancy (E) have a higher risk of mortality (O) than preterm infants with no history of exposure (C)?

### Sources and search strategy

A comprehensive literature search was undertaken using the PubMed and EMBASE from their inception up to March 2022. Search strategy is detailed in the supplementary material.

### Study selection

Studies were included if they examined extreme to moderate preterm infants(GA ≤34 weeks and/or birth weight ≤1500 gram) and reported primary data that could be used to measure the association between exposure to CA, HDP or SGA/IUGR and mortality before hospital discharge.

### Data extraction, definitions, and quality assessment

Two investigators (TH, EV-M) independently extracted data from relevant studies using a predetermined data extraction form. Any definition of chorioamnionitis, HDP or SGA/IUGR was accepted but we performed sub-group analysis based on the different definitions. Methodological quality was assessed using the Newcastle-Ottawa Scale (NOS) for cohort or case-control studies (28).

### Statistical analysis

Studies were combined and analyzed using COMPREHENSIVE META-ANALYSIS V3.0 software (Biostat Inc., Englewood, NJ, USA). Summary statistics were calculated with a random-effects model and subgroups were combined with a mixed-effects model (29). For dichotomous outcomes, the odds ratio (OR) with 95% confidence interval (CI) was calculated. For continuous outcomes, the mean difference (MD) with 95% CI was calculated. Statistical heterogeneity was assessed by Cochran’s Q statistic and by the I^2^ statistic. Potential sources of heterogeneity were assessed through subgroup analysis and/or random effects (method of moments), univariate meta-regression analysis (30). We used the Egger’s regression test and funnel plots to assess publication bias. A probability value of less than 0.05 (0.10 for heterogeneity) was considered statistically significant.

## Results

The PRISMA flow diagram of the search process is shown in eFigure 1 in the Supplement. Of 4322 potentially relevant studies, 150 met the inclusion criteria. These studies included 612,580 infants. Characteristics of the studies and quality assessment are summarized in eTable 1 in the Supplement. All studies received a NOS score of at least six points, indicating a low to moderate risk of bias.

**Fig. 1.**
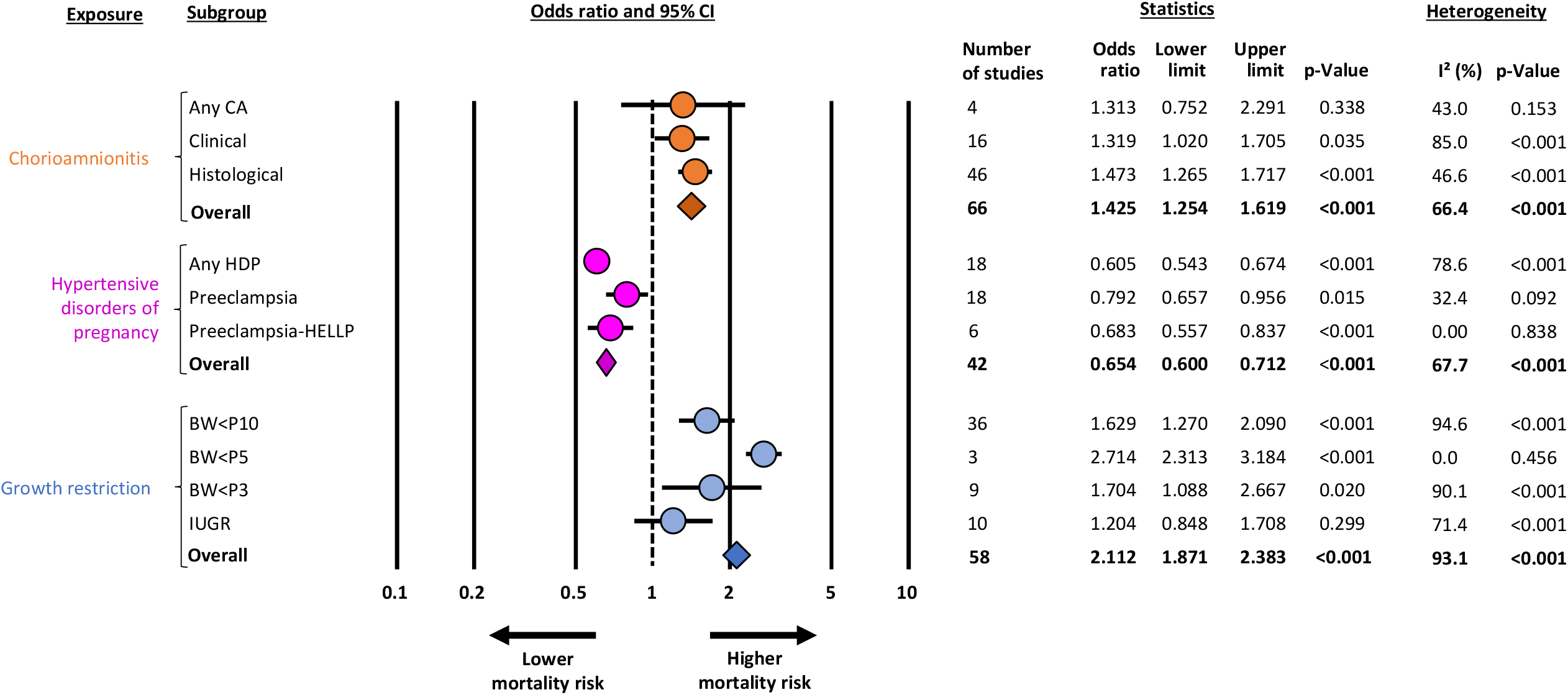
Summary of meta-analyses on the association between endotype of prematurity and mortality in infants up to 34 weeks of gestational age. BW: birth weight; CA: chorioamnionitis; CI: confidence interval; HDP: hypertensive disorders of pregnancy; HELLP: syndrome in pregnancy characterized by hemolysis, elevated liver enzymes and low platelet count; P3: 3^rd^ percentile; P5: 5^th^ percentile; P10: 10^th^ percentile; IUGR: intra-uterine growth restriction defined on the basis of fetal growth assessment

**Table 1.** Meta-analyses of other covariates.

| Exposure | Meta-analysis | K | Effect size | 95% CI |  | P | Heterogeneity |  |
| --- | --- | --- | --- | --- | --- | --- | --- | --- |
|  |  |  |  | Lower limit | Upper limit |  | I<sup>2</sup> (%) | P |
| CA | GA (MD in weeks) | 52 | -0.696 | -0.892 | -0.499 | <0.001 | 98.4 | <0.001 |
|  | Male sex (OR) | 41 | 0.969 | 0.889 | 1.056 | 0.474 | 52.8 | <0.001 |
|  | HDP (OR) | 21 | 0.110 | 0.076 | 0.160 | <0.001 | 81.3 | <0.001 |
|  | SGA/IUGR (OR) | 19 | 0.351 | 0.244 | 0.506 | <0.001 | 80.5 | <0.001 |
| HDP | GA (MD in weeks) | 28 | 0.271 | 0.187 | 0.355 | <0.001 | 97.9 | <0.001 |
|  | Male sex (OR) | 27 | 0.778 | 0.744 | 0.814 | <0.001 | 47.2 | 0.004 |
|  | CA (OR) | 12 | 0.132 | 0.080 | 0.219 | <0.001 | 89.0 | <0.001 |
|  | SGA/IUGR (OR) | 24 | 4.695 | 3.572 | 6.172 | <0.001 | 98.1 | <0.001 |
| SGA/IUGR | GA (MD in weeks) | 30 | 0.185 | 0.023 | 0.348 | 0.025 | 97.3 | <0.001 |
|  | Male sex (OR) | 31 | 0.950 | 0.851 | 1.060 | 0.355 | 79.2 | <0.001 |
|  | CA (OR) | 5 | 0.226 | 0.140 | 0.364 | <0.001 | 0.0 | 0.498 |
|  | HDP (OR) | 17 | 4.623 | 2.620 | 8.159 | <0.001 | 98.4 | <0.001 |

CA: chorioamnionitis; CI: confidence interval; GA: gestational age; HDP: hypertensive disorders of pregnancy; IUGR: intra-uterine growth retardation defined on the basis of fetal growth assessment; K: number of studies; MD: mean difference (exposed minus unexposed); OR: odds ratio; SGA: small for gestational age as defined by birth weight percentile.

### Main meta-analyses

Meta-analysis showed a significant association between chorioamnionitis and increased odds of mortality (Fig. 1, eFig. 2 in the Supplement). Heterogeneity was moderate (Fig. 1). When subdividing by chorioamnionitis definition, the association with mortality remained significant for both clinical and histological chorioamnionitis. Meta-regression did not show significant differences depending on choriomanionitis definition (P=0.375). When the meta-analysis was limited to the subgroup of infants with GA≤28 weeks, no significant association between choriomanonitis and mortality was observed (Fig. 2, eFig. 3 in the Supplement). Meta-regression showed significant differences (P=0.021) between studies that only included infants with GA≤28 weeks and those that also included older infants (eTable 2 in the Supplement).

**Table 2.** Subgroup analysis based on characteristics of the studies.

| Analysis | Criteria for subgrouping |  | K | OR | 95% CI |  | P | Heterogeneity |  | Meta-regression |  |
| --- | --- | --- | --- | --- | --- | --- | --- | --- | --- | --- | --- |
|  |  |  |  |  | Lower limit | Upper limit |  | I<sup>2</sup>(%) | P | P | R<sup>2</sup> analog |
| CA | Study type | Case-control | 5 | 2.059 | 1.181 | 3.589 | 0.011 | 26.5 | 0.245 | 0.181 | 0.00 |
|  |  | Cohort | 61 | 1.394 | 1.222 | 1.591 | <0.001 | 67.8 | <0.001 |  |  |
|  |  | Prospective | 41 | 1.404 | 1.187 | 1.662 | <0.001 | 55.3 | <0.001 | 0.975 | 0.00 |
|  |  | Retrospective | 25 | 1.450 | 1.182 | 1.778 | <0.001 | 76.5 | <0.001 |  |  |
|  | Maximum GA | GA≤28 | 13 | 1.135 | 0.894 | 1.441 | 0.297 | 70.4 | <0.001 | 0.017 | 0.33 |
|  |  | GA>28 | 53 | 1.522 | 1.340 | 1.730 | <0.001 | 47.1 | <0.001 |  |  |
|  | Year of study start | ≤2000 | 21 | 1.304 | 1.011 | 1.682 | 0.041 | 59.5 | <0.001 | 0.300 | 0.01 |
|  |  | >2000 | 45 | 1.485 | 1.278 | 1.725 | <0.001 | 68.1 | <0.001 |  |  |
| HDP | Study type | Case-control | 6 | 1.156 | 0.703 | 1.901 | 0.567 | 0.00 | 0.441 | 0.030 | 0.06 |
|  |  | Cohort | 36 | 0.644 | 0.588 | 0.705 | <0.001 | 69.8 | <0.001 |  |  |
|  |  | Prospective | 18 | 0.686 | 0.553 | 0.850 | 0.001 | 73.6 | <0.001 | 0.700 | 0.00 |
|  |  | Retrospective | 24 | 0.653 | 0.591 | 0.722 | <0.001 | 63.2 | <0.001 |  |  |
|  | Maximum GA | GA≤28 | 11 | 1.106 | 0.806 | 1.518 | 0.532 | 62.9 | 0.003 | <0.001 | 0.29 |
|  |  | GA>28 | 33 | 0.618 | 0.565 | 0.676 | <0.001 | 66.1 | <0.001 |  |  |
|  | Year of study start | ≤2000 | 20 | 0.669 | 0.571 | 0.784 | <0.001 | 67.6 | <0.001 | 0.606 | 0.00 |
|  |  | >2000 | 23 | 0.716 | 0.617 | 0.832 | <0.001 | 77.2 | <0.001 |  |  |
| SGA/IUGR | Study type | Case-control | 5 | 1.683 | 1.132 | 2.500 | 0.010 | 37.0 | 0.175 | 0.865 | 0.00 |
|  |  | Cohort | 53 | 1.609 | 1.324 | 1.957 | <0.001 | 93.6 | <0.001 |  |  |
|  |  | Prospective | 19 | 1.476 | 0.906 | 2.404 | 0.118 | 96.9 | <0.001 | 0.406 | 0.00 |
|  |  | Retrospective | 39 | 1.723 | 1.480 | 2.006 | <0.001 | 82.9 | <0.001 |  |  |
|  | Maximum GA | GA≤28 | 15 | 2.341 | 1.813 | 3.022 | <0.001 | 81.6 | <0.001 | 0.055 | 0.04 |
|  |  | GA>28 | 47 | 1.560 | 1.242 | 1.958 | <0.001 | 93.7 | <0.001 |  |  |
|  | Year of study start | ≤2000 | 32 | 1.462 | 1.100 | 1.942 | 0.009 | 94.4 | <0.001 | 0.274 | 0.01 |
|  |  | >2000 | 26 | 1.786 | 1.433 | 2.226 | <0.001 | 88.8 | <0.001 |  |  |

CA: chorioamnionitis; HDP: hypertensive disorders of pregnancy. SGA: small for gestational age as defined by birth weight percentile; IUGR: intra-uterine growth restriction defined on the basis of fetal growth assessment; GA: gestational age; wks: weeks; K: number of studies; OR: odds ratio; CI: confidence interval; P: p-value; I<sup>2</sup>: percentage of variation across studies that is due to heterogeneity; R<sup>2</sup> analog: total between-study variance explained by the moderator.

**Fig. 2.**
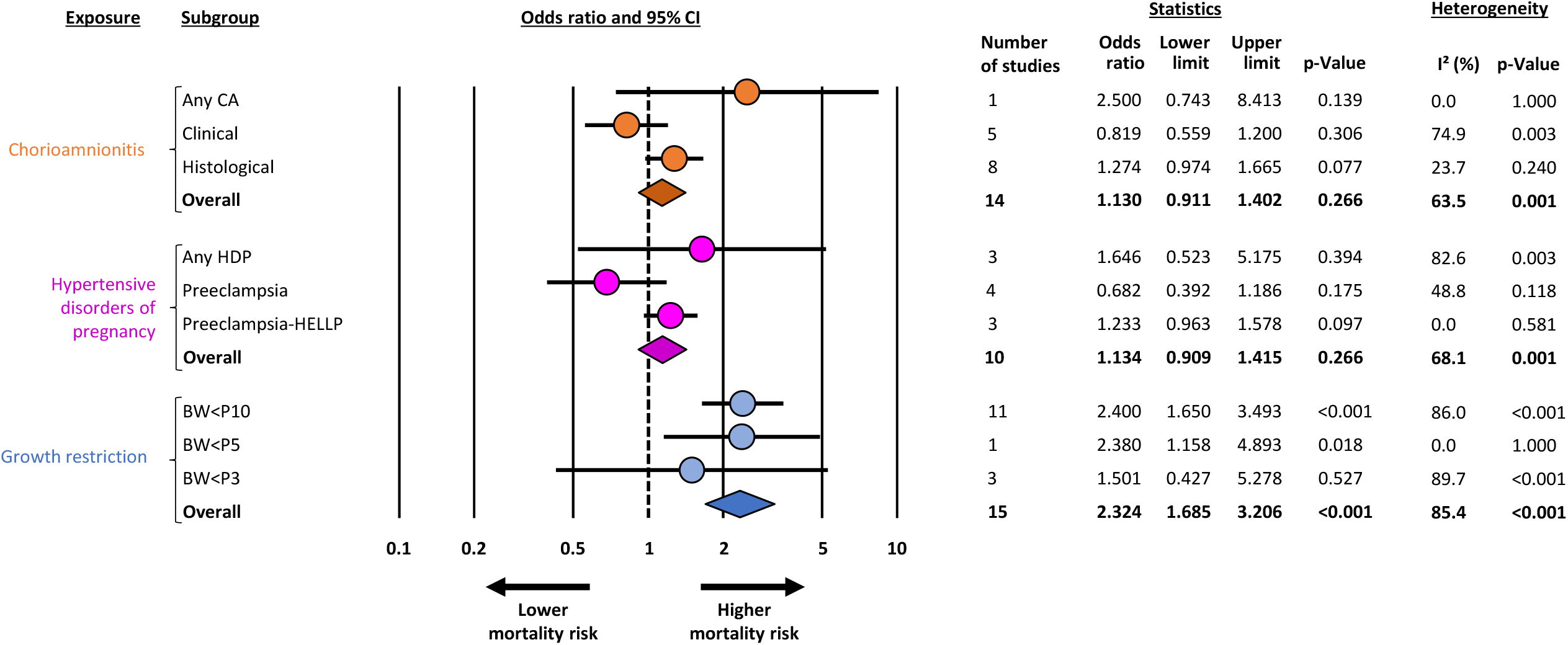
Summary of meta-analyses on the association between endotype of prematurity and mortality in infants up to 28 weeks of gestational age. BW: birth weight; CA: chorioamnionitis; CI: confidence interval; HDP: hypertensive disorders of pregnancy; HELLP: syndrome in pregnancy characterized by hemolysis, elevated liver enzymes and low platelet count; P3: 3rd percentile; P5: 5th percentile; P10: 10th percentile

**Fig. 3.**
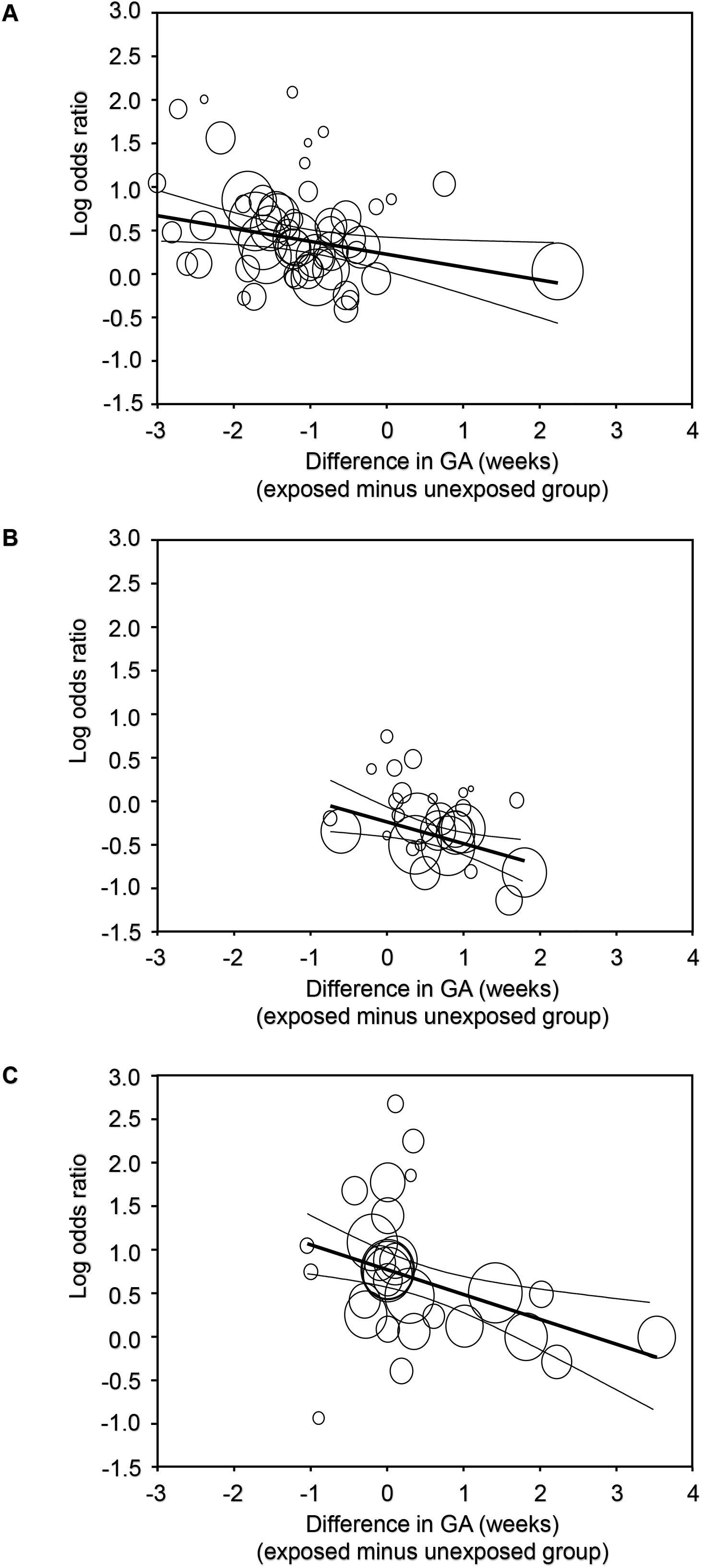
Meta-regression plot showing the correlation between the effect size of the association endotype-mortality and the mean difference (MD) in gestational age (GA) between exposed and non-exposed groups. (A) Univariate regression model correlating the difference in GA between chorioamnionitis-exposed and chorioamnionitis-unexposed infants. A total of 53 studies were included (coefficient -0.178; 95% CI -0.298 to -0.068; p = 0.002; R2 analog 0.37). Each week that chorioamnionitis-exposed infants were born earlier than control infants resulted in an increase in mortality log OR of 0.178 (the equivalent of going from an OR of 1.00 to an OR of 1.19). (B) Univariate regression model correlating the difference in GA between hypertensive disorders of pregnancy (HDP)-exposed and HDP-unexposed infants. A total of 32 studies were included (coefficient -0.249; 95% CI -0.403 to -0.094; p = 0.002; R2 analog 0.17). (C) Univariate regression model correlating the difference in GA between small for GA (SGA)/intrauterine growth restriction (IUGR)-exposed and SGA/IUGR-unexposed infants. A total of 34 studies were included (coefficient - 0.282; 95% CI -0.437 to -0.129; p<0.001; R2 analog 0.38).

Meta-analysis showed a significant association between HDP and decreased odds of mortality (Fig. 1, eFig. 4 in the Supplement). Heterogeneity was moderate (Fig. 1). When subdividing by HDP definition, the association with mortality remained significant for any HDP and preeclampsia but not for preeclampsia/HELLP. Meta-regression showed a significant difference (P=0.013) between the ORs for any HDP and preeclampsia. When the meta-analysis was limited to the subgroup of infants with GA≤28 weeks, no significant association between HDP and mortality was observed (Fig. 2, eFig. 5 in the Supplement). Meta-regression showed significant differences (P<0.001) between studies that only included infants with GA≤28 weeks and those that also included older infants (eTable 2 in the Supplement).

Meta-analysis showed a significant association between SGA/IUGR and increased odds of mortality (Fig. 1, eFig. 6 in the Supplement). Heterogeneity was high (Fig. 1). When subdividing by definition of SGA/IUGR, the association with mortality remained significant for the three thresholds of birth weight (BW <P10, <P5, and <P3), but not for IUGR (Fig. 1). Meta-regression analysis did not show significant differences in the association between SGA/IUGR and mortality depending on the criteria used to define SGA/IUGR (P=0.783). The association between SGA/IUGR and increased odds of mortality was also observed when the meta-analysis was limited to the subgroup of infants with GA≤28 weeks (Fig. 2, eFig. 7 in the Supplement).

Neither visual inspection nor Egger’s test suggested the presence of publication or selection bias for the chorioamnionitis (P=0.394) and SGA/IUGR (P=0.554) meta-analyses (eFig. 8 in the Supplement). In contrast, a slight publication bias was observed in the HDP meta-analysis (P=0.016).

### Additional meta-analyses, sensitivity analysis and meta-regression

To investigate the potential sources of heterogeneity, we conducted additional meta-analyses exploring the differences in baseline characteristics (GA, sex, and exposure to antenatal corticosteroids) between the groups exposed and non-exposed to chorioamnionitis, HDP, or SGA/IUGR. We also performed meta-regression and subgroup analysis.

Infants exposed to chorioamnionitis had a significantly lower GA than infants not exposed to the condition, while infants exposed to HDP or belonging to the SGA/IUGR group had a higher GA than their respective controls (Table 1). Meta-regression showed that these differences in GA significantly correlated with the effect size of the association between mortality and chorioamnionitis, HDP, or SGA/IUGR (Fig. 3, eTable 3 in the Supplement).

**Table 3.**
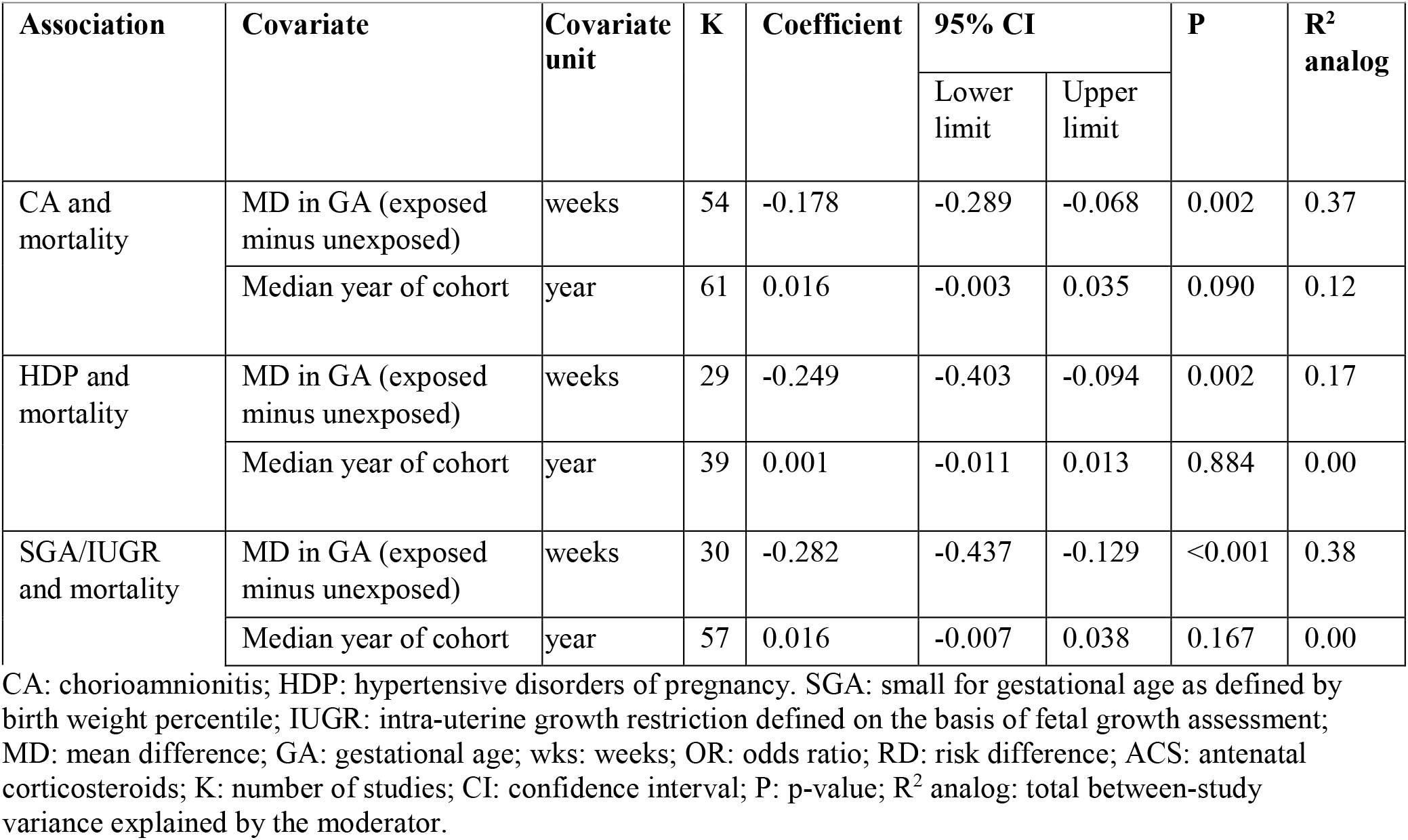
Meta-regression on continuous variables.

| Association | Covariate | Covariate unit | K | Coefficient | 95% CI |  | P | R<sup>2</sup> analog |
| --- | --- | --- | --- | --- | --- | --- | --- | --- |
|  |  |  |  |  | Lower limit | Upper limit |  |  |
| CA and mortality | MD in GA (exposed minus unexposed) | weeks | 54 | -0.178 | -0.289 | -0.068 | 0.002 | 0.37 |
|  | Median year of cohort | year | 61 | 0.016 | -0.003 | 0.035 | 0.090 | 0.12 |
| HDP and mortality | MD in GA (exposed minus unexposed) | weeks | 29 | -0.249 | -0.403 | -0.094 | 0.002 | 0.17 |
|  | Median year of cohort | year | 39 | 0.001 | -0.011 | 0.013 | 0.884 | 0.00 |
| SGA/IUGR and mortality | MD in GA (exposed minus unexposed) | weeks | 30 | -0.282 | -0.437 | -0.129 | <0.001 | 0.38 |
|  | Median year of cohort | year | 57 | 0.016 | -0.007 | 0.038 | 0.167 | 0.00 |

CA: chorioamnionitis; HDP: hypertensive disorders of pregnancy. SGA: small for gestational age as defined by birth weight percentile; IUGR: intra-uterine growth restriction defined on the basis of fetal growth assessment; MD: mean difference; GA: gestational age; wks: weeks; OR: odds ratio; RD: risk difference; ACS: antenatal corticosteroids; K: number of studies; CI: confidence interval; P: p-value; R<sup>2</sup> analog: total between-study variance explained by the moderator.

To further assess the effect of GA on the meta-analyses, we carried out a subgroup analysis of studies where the difference in GA between infants exposed to the chorioamnionitis, HDP, or SGA/IUGR and their respective controls was not statistically significant (p>0.05). In these subgroups of studies, neither chorioamnionitis nor HDP were significantly associated with mortality odds, but the association between SGA/IUGR and higher odds of mortality was maintained (eTable 4 in the Supplement).

**Table 4.** Subgroup analysis on gestational age.

| Analysis | Subgrouping on gestational age (GA) | K | OR | 95% CI |  | P | Heterogeneity |  | Meta-regression |  |
| --- | --- | --- | --- | --- | --- | --- | --- | --- | --- | --- |
|  |  |  |  | Lower limit | Upper limit |  | I<sup>2</sup>(%) | P | P | R<sup>2</sup> analog |
| CA | GA significantly lower in CA group | 41 | 1.471 | 1.281 | 1.689 | <0.001 | 61.7 | <0.001 | 0.432 | 0.11 |
|  | GA not significantly different | 9 | 1.257 | 0.951 | 1.662 | 0.109 | 0.00 | 0.897 |  |  |
| HDP | GA significantly higher in HDP group | 15 | 0.611 | 0.534 | 0.698 | <0.001 | 62.7 | 0.001 | 0.014 | 0.00 |
|  | GA not significantly different | 12 | 0.884 | 0.690 | 1.132 | 0.329 | 65.7 | 0.001 |  |  |
| SGA/IUGR | GA significantly higher in SGA/IUGR group | 7 | 1.245 | 0.950 | 1.632 | 0.112 | 48.8 | 0.069 | 0.025 | 0.00 |
|  | GA not significantly different | 19 | 2.383 | 1.965 | 2.889 | <0.001 | 65.0 | <0.001 |  |  |

CA: chorioamnionitis; HDP: hypertensive disorders of pregnancy. SGA: small for gestational age as defined by birth weight percentile; IUGR: intra-uterine growth restriction defined on the basis of fetal growth assessment; GA: gestational age; wks: weeks; K: number of studies; OR: odds ratio; CI: confidence interval; P: p-value; I<sup>2</sup>: percentage of variation across studies that is due to heterogeneity; R<sup>2</sup> analog: total between-study variance explained by the moderator.

Chorioamnionitis, HDP and SGA/IUGR were all associated with a higher odds of exposure to antenatal corticosteroids compared to their respective control groups (eTable 6 in the Supplement). Exposure to HDP was negatively associated with male sex (Table 1). As expected, exposure to chorioamnionitis was negatively associated with exposure to HDP or being SGA/IUGR and a strong positive association was observed between HDP and SGA (Table 1).

**Table 5.** Subgroup analysis for continent.

| Analysis | Continent | K | OR | 95% CI |  | P | Heterogeneity |  | Meta-regression |  |
| --- | --- | --- | --- | --- | --- | --- | --- | --- | --- | --- |
|  |  |  |  | Lower limit | Upper limit |  | I<sup>2</sup>(%) | P | P | R<sup>2</sup> analog |
| CA | Asia | 14 | 1.163 | 0.892 | 1.515 | 0.026 | 47.1 | 0.012 | 0.061 | 0.20 |
|  | Oceania | 5 | 1.381 | 1.126 | 1.694 | 0.002 | 0.00 | 0.717 | 0.581 | 0.00 |
|  | Europe | 31 | 1.601 | 1.294 | 1.981 | <0.001 | 70.1 | <0.001 | Ref. |  |
|  | North America | 16 | 1.368 | 1.127 | 1.660 | 0.002 | 47.0 | 0.020 | 0.356 | 0.03 |
| HDP | Asia | 4 | 0.753 | 0.641 | 0.884 | 0.001 | 0.00 | 0.820 | 0.123 | 0.04 |
|  | Oceania | 2 | 0.645 | 0.474 | 0.878 | 0.005 | 0.00 | 0.913 | 0.818 | 0.00 |
|  | Europe | 15 | 0.635 | 0.516 | 0.781 | <0.001 | 66.8 | <0.001 | Ref. |  |
|  | America | 19 | 0.633 | 0.555 | 0.721 | <0.001 | 69.7 | <0.001 | 0.608 | 0.00 |
| SGA/IUGR | Asia | 9 | 1.452 | 1.039 | 2.029 | 0.029 | 87.1 | <0.001 | 0.330 | 0.00 |
|  | Oceania | 5 | 2.204 | 1.065 | 4.561 | 0.033 | 93.6 | <0.001 | 0.722 | 0.00 |
|  | Europe | 23 | 1.867 | 1.479 | 2.357 | <0.001 | 78.2 | <0.001 | Ref. |  |
|  | North America | 19 | 1.473 | 1.025 | 2.117 | 0.036 | 96.3 | <0.001 | 0.222 | 0.00 |

CA: chorioamnionitis; HDP: hypertensive disorders of pregnancy. SGA: small for gestational age as defined by birth weight percentile; IUGR: intra-uterine growth retardation defined on the basis of fetal growth assessment; GA: gestational age; wks: weeks; K: number of studies; OR: odds ratio; CI: confidence interval; P: p-value; I<sup>2</sup>: percentage of variation across studies that is due to heterogeneity. Ref.: reference; R<sup>2</sup> analog: total between-study variance explained by the moderator.

**Table 6.** Meta-analysis on antenatal corticosteroids.

| Analysis | Exposure to antenatal corticosteroids (ACS) | K | OR | 95% CI |  | P | Heterogeneity |  |
| --- | --- | --- | --- | --- | --- | --- | --- | --- |
|  |  |  |  | Lower limit | Upper limit |  | I<sup>2</sup>(%) | P |
| CA | Undefined | 27 | 1.282 | 1.091 | 1.505 | 0.005 | 68.3 | <0.001 |
|  | Any | 8 | 1.112 | 0.806 | 1.535 | 0.518 | 75.5 | <0.001 |
|  | Complete course | 7 | 1.042 | 0.795 | 1.366 | 0.764 | 45.4 | 0.089 |
| HDP | Undefined | 10 | 1.677 | 1.382 | 2.034 | <0.001 | 69.0 | 0.001 |
|  | Any | 10 | 1.238 | 0.761 | 2.017 | 0.390 | 98.7 | <0.001 |
|  | Complete course | 5 | 1.847 | 1.232 | 2.770 | 0.003 | 69.4 | 0.011 |
| SGA/IUGR | Undefined | 16 | 1.258 | 1.101 | 1.438 | 0.001 | 65.3 | <0.001 |
|  | Any | 4 | 1.148 | 0.960 | 1.372 | 0.131 | 47.3 | 0.128 |
|  | Complete course | 9 | 1.381 | 0.993 | 1.920 | 0.055 | 88.5 | <0.001 |

CA: chorioamnionitis; HDP: hypertensive disorders of pregnancy. SGA: small for gestational age as defined by birth weight percentile; IUGR: intra-uterine growth retardation defined on the basis of fetal growth assessment; wks: weeks; K: number of studies; OR: odds ratio; CI: confidence interval; P: p-value; I<sup>2</sup>: percentage of variation across studies that is due to heterogeneity.

We also analyzed the possible effect of study design, median year in which the study was conducted, and geographical location (continent) on the results of the meta-analysis. Meta-regression showed no significant differences when studies were grouped based on whether they were prospective or retrospective (eTable 2 in the Supplement). When cohort and case-control studies were compared, a significant difference was observed for the HDP meta-analysis (P=0.030), but not for the meta-analyses on chorioamnionitis or SGA/IUGR (eTable 2 in the Supplement). Meta-regression did not show a significant correlation between the median year in which the studies were conducted and the effect size of the association between mortality and chorioamnionitis, HDP, or SGA/IUGR (eTable 3 in the Supplement). In addition, meta-regression also showed no significant differences when studies were grouped according to whether the inclusion of infants took place before or after the year 2000 (eTable 2 in the Supplement). Finally, meta-regression showed no significant differences when the studies were grouped by continent (eTable 5 in the Supplement).

## Discussion/Conclusion

This is the first systematic review and meta-analysis that comprehensively addresses the association between endotype of prematurity and mortality during first hospital admission. Meta-analysis showed an association between the inflammatory/infectious endotype, represented by chorioamnionitis, and increased odds of mortality. In contrast, meta-analysis of the two proxies of the placental dysfunction endotype showed diametrically opposite results; HDP were associated with decreased odds while SGA/IUGR was associated with positive odds of mortality. Interestingly, when analysis was restricted to extremely preterm infants, only the association between SGA/IUGR and mortality remained significant. In addition, our results confirm that conditions triggering prematurity differ in important characteristics that may play a relevant role in mortality risk. Chorioamnionitis was associated with lower GA and higher exposure to antenatal corticosteroids, while HDP and SGA/IUGR were associated with higher GA. Meta-regression demonstrated a significant correlation between these differences in GA and the effect size of the association between mortality and endotype. Meta-regression also showed that the association between endotype of prematurity and mortality is relatively homogeneous across continents and has not changed significantly over the years.

Since very and extremely preterm birth is by definition a pathological condition, there is no healthy control group with which to compare infants exposed to a given prenatal insult (4, 5, 9, 31). Although some placentas may combine lesions of infection/inflammation and vascular dysfunction (32), fetuses exposed to chorioamnionitis will be less likely exposed to HDP and *vice versa* (4, 5, 9, 31). The main originality of our meta-analysis lies in the comparison of the three pathological conditions most frequently associated with preterm birth. However, both our identification of the two endotypes and the validity of chorioamnionitis, HDP and SGA/IUGR as their proxies may be questionable. Moreover, there is great heterogeneity in the potential clinical applicability of these three proxies. For example, histological chorioamnionitis may be a good indicator of the presence of the infectious endotype, but information about its presence may be not available at birth (8).

Regarding the endotype of placental dysfunction, the criteria for the diagnosis of HDP have been changing over the years and even those on which there is greater consensus may be far from being implemented worldwide (33). Finally, the most questionable assumption of our study may be the use of SGA as proxy for placental dysfunction. SGA is a statistical definition based on BW, with the 10^th^ percentile as the most commonly used threshold (26, 34, 35). Therefore, SGA definition also encompasses constitutionally small infants who do not suffer growth restriction. Conversely, growth restricted infants who have a BW above the 10^th^ percentile may be misclassified as normally grown (26). It has been suggested that a cutoff value below the 5th or 3rd percentile may reflect a degree of smallness that is more likely to be pathologic rather than constitutional (26, 36). However, even this pathology may be of fetal origin as there are conditions in which restricted growth is not caused primarily by placental insufficiency, but indirectly leads to it (37).

Regardless of its specificity as a marker of placental dysfunction, our data confirmed that SGA preterm infants are at increased risk of death. The association between mortality and SGA was observed even for the most conservative threshold (10^th^ percentile). Preterm SGA infants add to their immaturity the technical challenges of their small size that complicate procedures such as endotracheal intubation or vascular access (38, 39). Of note, the meta-analysis could not demonstrate a significant association between mortality and growth restriction based on fetal assessment. However, this analysis was based on very few studies that showed a high degree of heterogeneity. Damodaram et al. conducted a meta-analysis in which cohorts of growth-restricted preterm infants were compared with cohorts of infants with normal growth and found a marked increase in mortality across all GAs (19).

A common conundrum in perinatal medicine is the extent to which complications of pregnancy harm preterm infants through triggering prematurity or through disturbances in fetal homeostasis and development (4, 40). In the last few years, we conducted meta-analyses on the association between chorioamnionitis and complications of prematurity such as bronchopulmonary dysplasia (23), retinopathy of prematurity (41), intraventricular hemorrhage (42), sepsis (43), or patent ductus arteriosus (44). Similar to the present findings, meta-analyses showed an association between chorioamnionitis and all these conditions (23, 41-44). In addition, meta-regression analysis showed a correlation between the earlier GA of the chorioamnionitis-exposed infants and the risk of developing the above-mentioned complications (23, 41-44). These data suggest that a significant component of the pathologic effect of chorioamnionitis is related to lower GA of the infants rather than to the inflammatory stress that generates in the fetus.

In contrast to what we observed for chorioamnionitis and for SGA/IUGR, exposure to HDP was associated with lower odds of mortality in the newborn. Placental dysfunction is a multifactorial condition, with fetuses becoming hypoxemic, undernourished, and hypercortisolemic, each of these variables being capable of independently affecting fetal homeostasis and development (45). Our results suggest that when placental dysfunction does not significantly affect birth weight, preterm newborns may be in more favorable conditions to cope with extrauterine life. However, it cannot be overlooked that HDP-exposed infants had a higher GA and were more often girls, which may have contributed to the lower mortality (46).

Even among infants of comparable GA at delivery, the endotype of prematurity may be a determining factor in the time and cause of death. Most deaths in extreme preterm infants occur in the first days of life as a consequence of pulmonary immaturity (6, 47). It is noteworthy that the intrauterine stresses associated with both the infectious/inflammatory endotype and the placental dysfunction endotype have been considered as inducers of pulmonary maturity, leading to a lower rate of respiratory distress at birth (48, 49). Although very limited by the clinical and statistical heterogeneity, the results of our previous meta-analyses suggest that the lower incidence of respiratory distress is observed only in the SGA/IUGR group (7, 23). Nevertheless, other conditions such as pulmonary hemorrhage or pulmonary hypertension may contribute to increased early respiratory mortality in SGA infants (6, 47).

Severe intraventricular hemorrhage (IVH) is another major cause of early mortality among extremely preterm infants (6, 47, 50). As mentioned above, in a previous meta-analysis we observed an association between chorioamnionitis and the risk of developing IVH (42). Interestingly, this association was independent on the effect of chorioamnionitis on GA (42). It is therefore likely that a part of the early mortality induced by chorioamnionitis is related to clinical instability that may increase the chances of developing severe IVH. Conversely, and although to our knowledge this has not been systematically reviewed, several cohort studies suggest that exposure to HDP or being SGA may be associated with a lower risk of developing severe IVH (6, 47, 51, 52).

An important limitation of our study is the moderate to high heterogeneity that we found in most of the meta-analyses. Through subgroup analysis and meta-regression, we aimed to investigate possible sources of heterogeneity. Neither the association between chorioamnionitis and high odds of mortality nor the association HDP and reduced odds of mortality were observed in the subgroup of studies that exclusively included extremely preterm infants (GA ≤28 weeks). In contrast, the association between SGA/IUGR and high odds of mortality was maintained in the subgroup of extremely preterm infants. Together, these data suggest that at lower GAs mortality is relatively independent of the endotype of prematurity. However, infants who combine extreme prematurity and being SGA would have the highest mortality risk.

Another limitation of our meta-analyses is that they are based on a ‘births-based’ instead of ‘fetuses-at-risk’ formulation (53, 54). Fetuses who would be viable but die *in utero* are not considered when assessing mortality from a births-based perspective (53, 54). Therefore, if we intend to analyze the overall impact of an endotype of prematurity on perinatal health, we should also consider how many children are not born prematurely because they die as a result of the severely adverse intrauterine environment that is induced by their particular endotype.

Advances in perinatology over the past two decades, such as wide implementation of antenatal steroids and surfactant, have changed care for extremely preterm infants and have been associated with a decrease in mortality (55). However, neither meta-regression nor division of studies into two subgroups (median year of the cohort before or after 2000) could demonstrate an effect of study date on the association between endotype of prematurity and mortality. Finally, it is interesting to note our finding that in the three groups analyzed (chorioamnionitis, HDP and SGA/IUGR) the odds of receiving antenatal corticosteroids was higher than that of the respective controls. We speculate that this finding may be related to the more heterogeneous nature of the control groups. Nevertheless, our group is conducting a meta-analysis on the association between antenatal corticosteroids and endotype of prematurity.

In summary, the present data suggest that the infectious/inflammatory endotype of prematurity has a greater overall impact on mortality risk as it is the most frequent endotype in the lower and more vulnerable gestational ages. However, when the endotype of placental dysfunction is severe enough to induce growth restriction, it is strongly associated with higher mortality rates even though newborns are more mature. Nevertheless, neither of the two endotypes is desirable and extremely preterm birth should be prevented regardless of the etiopathogenic process that caused it. However, delay in delivery may not benefit some infants if prematurity is triggered by a severe pathology and postponement of delivery is not adequately addressing the underlying condition (15). Finally, since some placental diagnoses may be critical for personalized clinical care of extremely preterm newborns, efforts should be made to have information on placental pathology available in the first days of life (8, 32).

## Statement of Ethics

As this systematic review and meta-analysis did not involve animal subjects or personally identifiable information on human subjects, ethics review board approval and patient consent were not required.

## Data Availability

All data produced in the present study are available upon reasonable request to the authors

## Author Contributions

EV conceived and designed the study with input from the other authors. TH and EV-M designed and executed the literature search, screened and reviewed the search results and abstracted the data. EV checked data extraction for accuracy and completeness. TH and EV-M assessed the quality of the included studies. TH conducted the analysis with input from EV. All authors contributed to the interpretation of analysis. TH and EV made the figures and tables. TH and EV drafted the manuscript with input from the other authors. All authors reviewed the manuscript and provided important intellectual content. TH and EV take responsibility for the article as a whole.

## Data Availability Statement

All data relevant to the study are included in the article or uploaded as supplementary information. Additional data are available upon reasonable request.

## Supplementary Online Content

## 1. Methods

### 1.1 Search strategy

**PubMed**

((Chorioamnionitis[Mesh] OR chorioamnionitis [TIAB] OR CA [TIAB] OR amnionitis [TIAB] OR funisitis [TIAB] OR fetal membranes, premature rupture [Mesh] OR placenta diseases [Mesh] OR placenta disease [TIAB] OR PPROM[TIAB] OR preterm premature rupture of membrane*[TIAB])

AND (mortality[Mesh] OR mortality, premature [Mesh] OR perinatal mortality [Mesh] OR infant mortality [Mesh] OR mortality[TIAB] OR death[TIAB] mortality[Mesh] OR mortality, premature [Mesh] OR perinatal mortality [Mesh] OR infant mortality [Mesh] OR mortality[TIAB] OR death[TIAB] OR short-term complication [TIAB] short term complication [TIAB] OR complication [TIAB])

AND (infant* [TIAB] OR Infants, Newborn[Mesh] OR Newborn*[TIAB] OR Neonate*[TIAB] OR neonatal[TIAB] OR Decreased gestational age [TIAB] OR premature gestation [TIAB] OR prematurity [TIAB] OR preterm [TIAB] OR premature [TIAB]))

**<u>OR</u>**

((Placental insufficiency [Mesh] OR placental insufficiency [TIAB] OR placental dysfunction [TIAB] OR Hypertension, pregnancy-induced [TIAB] OR pregnancy induced hypertension[TIAB] OR gestational hypertension[TIAB] OR pregnancy transient hypertension [TIAB] OR GHD [TIAB] OR pre-eclampsia [Mesh] OR pre-eclampsia [TIAB] OR preeclampsia [TIAB] OR HELLP syndrome [Mesh] OR HELLP syndrome [TIAB])

AND (mortality[Mesh] OR mortality, premature [Mesh] OR perinatal mortality [Mesh] OR infant mortality [Mesh] OR mortality[TIAB] OR death[TIAB] mortality[Mesh] OR mortality, premature [Mesh] OR perinatal mortality [Mesh] OR infant mortality [Mesh] OR mortality[TIAB] OR death[TIAB] OR short-term complication [TIAB] short term complication [TIAB] OR complication [TIAB])

AND (infant* [TIAB] OR Infants, Newborn[Mesh] OR Newborn*[TIAB] OR Neonate*[TIAB] OR neonatal[TIAB] OR Decreased gestational age [TIAB] OR premature gestation [TIAB] OR prematurity [TIAB] OR preterm [TIAB] OR premature [TIAB]))

**<u>OR</u>**

((Fetal growth restriction [Mesh] OR fetal growth restriction [TIAB] OR IUGR OR intrauterine growth restriction[TIAB] OR intrauterine growth restriction [TIAB] OR infant, small for gestational age [TIAB] OR SGA OR small for gestational age [TIAB])

AND (mortality[Mesh] OR mortality, premature [Mesh] OR perinatal mortality [Mesh] OR infant mortality [Mesh] OR mortality[TIAB] OR death[TIAB] mortality[Mesh] OR mortality, premature [Mesh] OR perinatal mortality [Mesh] OR infant mortality [Mesh] OR mortality[TIAB] OR death[TIAB] OR short-term complication [TIAB] short term complication [TIAB] OR complication [TIAB])

AND (infant* [TIAB] OR Infants, Newborn[Mesh] OR Newborn*[TIAB] OR Neonate*[TIAB] OR neonatal[TIAB] OR Decreased gestational age [TIAB] OR premature gestation [TIAB] OR prematurity [TIAB] OR preterm [TIAB] OR premature [TIAB]))

**Embase**

((exp chorioamnionitis/ OR chorioamnionitis.ti,ab. OR CA.ti,ab. OR amnionitis.ti,ab. OR funisitis.ti,ab. OR exp premature fetus membrane rupture/ OR exp placenta disorder/ OR placenta disease.ti,ab. OR PPROM.ti,ab. OR preterm premature rupture of membrane.ti,ab.) AND (exp mortality/ OR mortality.ti,ab. OR exp premature mortality/ OR premature mortality.ti,ab. OR exp perinatal mortality/ OR perinatal mortality.ti,ab. OR exp newborn mortality/ OR newborn mortality.ti,ab. OR exp infant mortality/ OR infant mortality.ti,ab. OR mortality.ti,ab. OR death.ti,ab. AND exp complication/ OR complication.ti,ab. OR short-term complication.ti,ab. OR short term complication.ti,ab.) AND (exp prematurity/ OR infant.ti,ab. OR preterm infant.ti,ab. OR newborn.ti,ab. OR neonate.ti,ab. OR neonatal.ti,ab. OR decreased gestational age.ti,ab. OR premature gestation.ti,ab. OR prematurity.ti,ab. OR preterm.ti,ab. OR premature.ti,ab.))

**<u>OR</u>**

((exp placental insufficiency/ OR placental insufficiency.ti,ab. OR placental dysfunction.ti,ab. OR exp maternal hypertension/ OR pregnancy induced hypertension.ti,ab. OR gestational hypertension.ti,ab. OR pregnancy transient hypertension.ti,ab. OR GHD.ti,ab. OR pre-eclampsia OR HELLP) AND (exp mortality/ OR mortality.ti,ab. OR exp premature mortality/ OR premature mortality.ti,ab. OR exp perinatal mortality/ OR perinatal mortality.ti,ab. OR exp newborn mortality/ OR newborn mortality.ti,ab. OR exp infant mortality/ OR infant mortality.ti,ab. OR mortality.ti,ab. OR death.ti,ab. AND exp complication/ OR complication.ti,ab. OR short-term complication.ti,ab. OR short term complication.ti,ab.) AND (exp prematurity/ OR infant.ti,ab. OR preterm infant.ti,ab. OR newborn.ti,ab. OR neonate.ti,ab. OR neonatal.ti,ab. OR decreased gestational age.ti,ab. OR premature gestation.ti,ab. OR prematurity.ti,ab. OR preterm.ti,ab. OR premature.ti,ab.))

**<u>OR</u>**

((exp intrauterine growth restriction/ OR fetal growth restriction.ti,ab. OR fetal growth restriction.ti,ab. OR IUGR.ti,ab. OR intrauterine growth restriction.ti,ab. OR intrauterine growth restriction.ti,ab. OR SGA.ti,ab. OR small for gestational age.ti,ab.) AND (exp mortality/ OR mortality.ti,ab. OR exp premature mortality/ OR premature mortality.ti,ab. OR exp perinatal mortality/ OR perinatal mortality.ti,ab. OR exp newborn mortality/ OR newborn mortality.ti,ab. OR exp infant mortality/ OR infant mortality.ti,ab. OR mortality.ti,ab. OR death.ti,ab. AND exp complication/ OR complication.ti,ab. OR short-term complication.ti,ab. OR short term complication.ti,ab.) AND (exp prematurity/ OR infant.ti,ab. OR preterm infant.ti,ab. OR newborn.ti,ab. OR neonate.ti,ab. OR neonatal.ti,ab. OR decreased gestational age.ti,ab. OR premature gestation.ti,ab. OR prematurity.ti,ab. OR preterm.ti,ab. OR premature.ti,ab.))

No language limits were set. Narrative reviews, systematic reviews, case reports, letters, editorials, and commentaries were excluded, but read to identify potential additional studies. Additional strategies to identify studies included manual review of reference lists from key articles that fulfilled our eligibility criteria, use of “related articles” feature in PubMed, and use of the “cited by” tool in Web of Science and Google scholar. Two reviewers independently screened the results of the searches, and included studies according to the inclusion criteria using EndNote (RRID:SCR_014001), using the methodology described by Bramer et al. (1)

### 1.2 Supplementary information on methods

#### Study selection

Studies were included if they examined preterm (GA <34 weeks) and/or very low birth weight (<1500g) infants and reported data that could be used to measure the association between exposure to chorioamnionitis, hypertensive disorders of pregnancy (HDP) or small for gestational age (SGA)/intrauterine growth restriction (IUGR) and mortality before hospital discharge. Therefore, we selected observational studies in which the exposure (chorioamnionitis, HDP or SGA/IUGR) was the independent variable and the outcome (mortality before hospital discharge) the dependent variable as well as studies in which the outcome was the independent variable and the exposure the dependent variable. Studies that exclusively included late preterm infants (GA ≥34 weeks) or that combined preterm and term infants were excluded. Due to the high number of included studies, no additional efforts were made to clarify definitions or other data with the authors. Abstracts and unpublished studies were also excluded. To identify relevant studies, two reviewers (TH, EV-M) independently screened the results of the searches and applied inclusion criteria using a structured form. Discrepancies were resolved through discussion or consultation with a third reviewer (EV).

#### Data extraction and quality assessment

Data extracted from each study included citation information, language of publication, country of conducted research, time period of the study, study objectives, study design, inclusion/exclusion criteria, definition criteria for chorioamnionitis, HDP, IUGR, SGA and mortality, patient characteristics, and results (including raw numbers or summary statistics when raw numbers were not available). A study was categorized as a case-control when each case was matched with a control in a predetermined ratio (1:1, 1:2, 1:3, etc.). A study was considered a cohort study when it included, prospectively or retrospectively, a group of infants during a given period, regardless of the number of infants included. Therefore, small cohorts were also included.

Any definition of chorioamnionitis, HDP, IUGR, or SGA was accepted but we performed sub-group analysis based on the different definitions. When a study used more than one definition criteria for growth restriction, definitions based on assessment of fetal growth (i.e., IUGR) prevailed over definitions based on BW. When a study used different BW threshold percentiles to define SGA, data from the lowest percentile were included. When a study did not specify the threshold percentile used, it was grouped together with the studies that used the 10th percentile.

Methodological quality was assessed using the Newcastle-Ottawa Scale for cohort or case-control studies.(2) This scale uses a star rating system (range: 0–9 stars) scoring three aspects of the study: selection (0–4), comparability (0–2) and exposure/outcome (0–3). Two reviewers (TH, EV-M) independently assessed the methodological quality of each study. Discrepancies were resolved through discussion.

#### Statistical analysis

Studies were combined and analyzed using COMPREHENSIVE META-ANALYSIS V3.0 software (Biostat Inc., Englewood, NJ, USA). For dichotomous outcomes, the odds ratio (OR) with 95% confidence interval (CI) was calculated from the data provided in the studies. Reported ORs were included when studies reported them and did not include the numerical data for the calculation of the OR. For continuous outcomes, the mean difference with 95% CI was calculated. When studies reported continuous variables as median and range or interquartile range, we estimated the mean and standard deviation using the method of Wan et al. and the calculator they provided. (3)

Due to anticipated heterogeneity, summary statistics were calculated with a random-effects model. This model accounts for variability between studies as well as within studies. Subgroup analyses were conducted according to the mixed-effects model.(4) In this model, a random-effects model is used to combine studies within each subgroup, and a fixed-effect model is used to combine subgroups and yield the overall effect. The study-to-study variance (tau-squared) is not assumed to be the same for all subgroups. This value is computed within subgroups and not pooled across subgroups. Statistical heterogeneity was assessed by Cochran’s *Q* statistic and by the *I*^*2*^ statistic, which is derived from *Q* and describes the proportion of total variation that is due to heterogeneity beyond chance.(5) The I^2^ statistic was interpreted as follows: low heterogeneity (25% ≤ I^2^ < 50%), moderate heterogeneity (50% ≤ I^2^ < 75%), and high heterogeneity (I^2^ ≥ 75%). We used the Egger’s regression test and funnel plots to assess publication bias. A probability value of less than 0.05 (0.10 for heterogeneity) was considered statistically significant.

Potential sources of heterogeneity were assessed through subgroup analysis and/or random effects (method of moments) univariate meta-regression analysis.^5^ For continuous covariates (examples: difference in mean gestational age between infants exposed and unexposed to chorioamnionitis) we used meta-regression analyses to test whether there was a significant relationship between the covariate and effect size, as indicated by a Z-value and an associated p-value. Meta-regression coefficient indicates the change in the log of the OR of the association between mortality and the corresponding exposure for a unit change in the predictor covariate.

Subgroups were compared using meta-regression for categorical covariates. For both categorical and continuous covariates, the R^2^ analog, defined as the total between-study variance explained by the moderator, was calculated based on the meta-regression matrix.(6) Covariates defined a priori were: 1) Continuous: study time (median year of cohort inclusion), differences between exposed and unexposed infants on GA; 2) Categorical: chorioamnionitis type, HDP type, SGA/IUGR type, mean difference of GA between exposed and unexposed infants statistically significant (P<0.05, yes/no), geographical location (continent) of the study, prospective vs retrospective and cohort vs case-control study. The clinical covariates were selected based on their relevance on the outcome of mortality.

## 2. Results

**eFigure 1.**
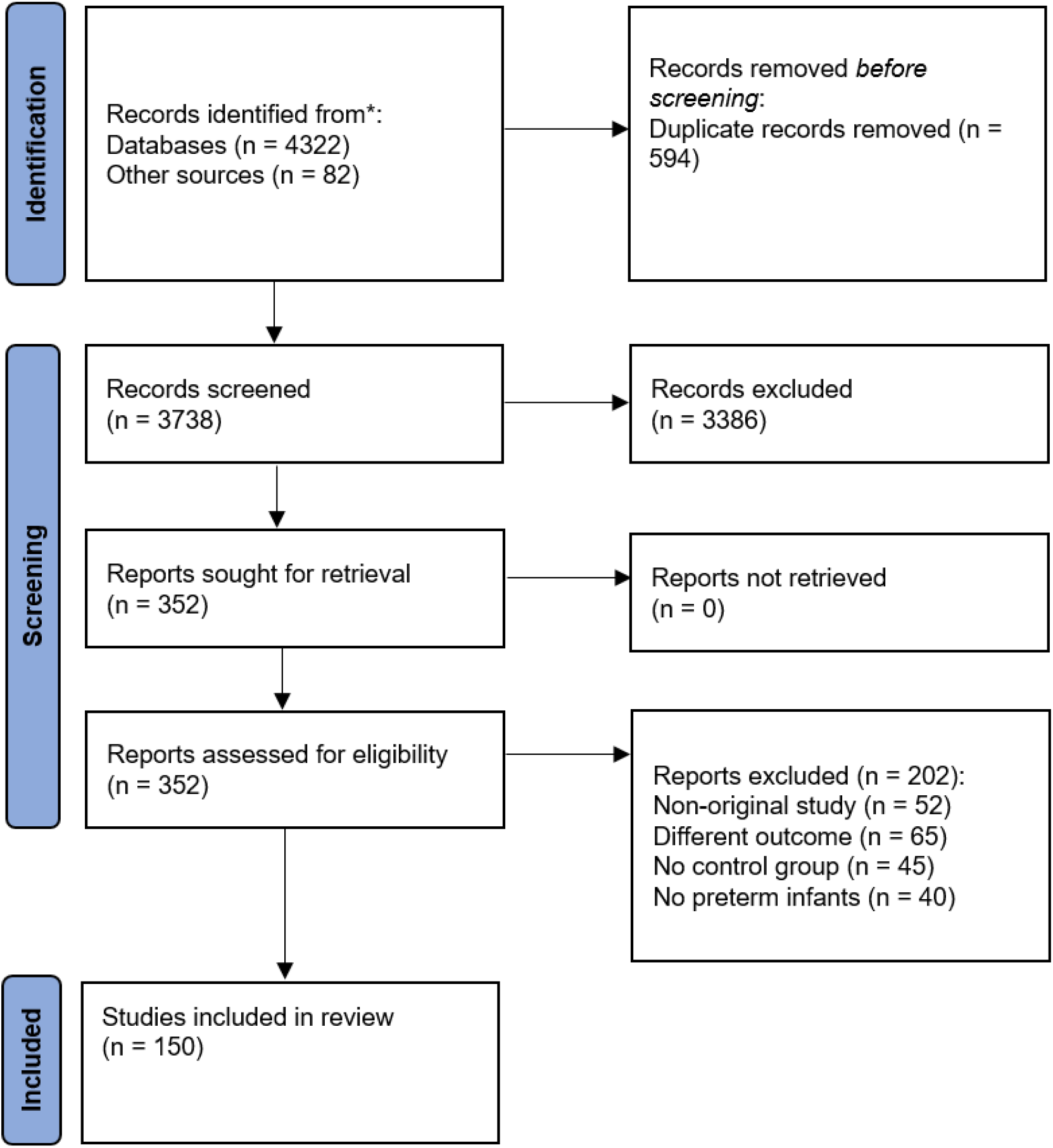
PRISMA Diagram of the Systematic Search.

**eFigure 2.**
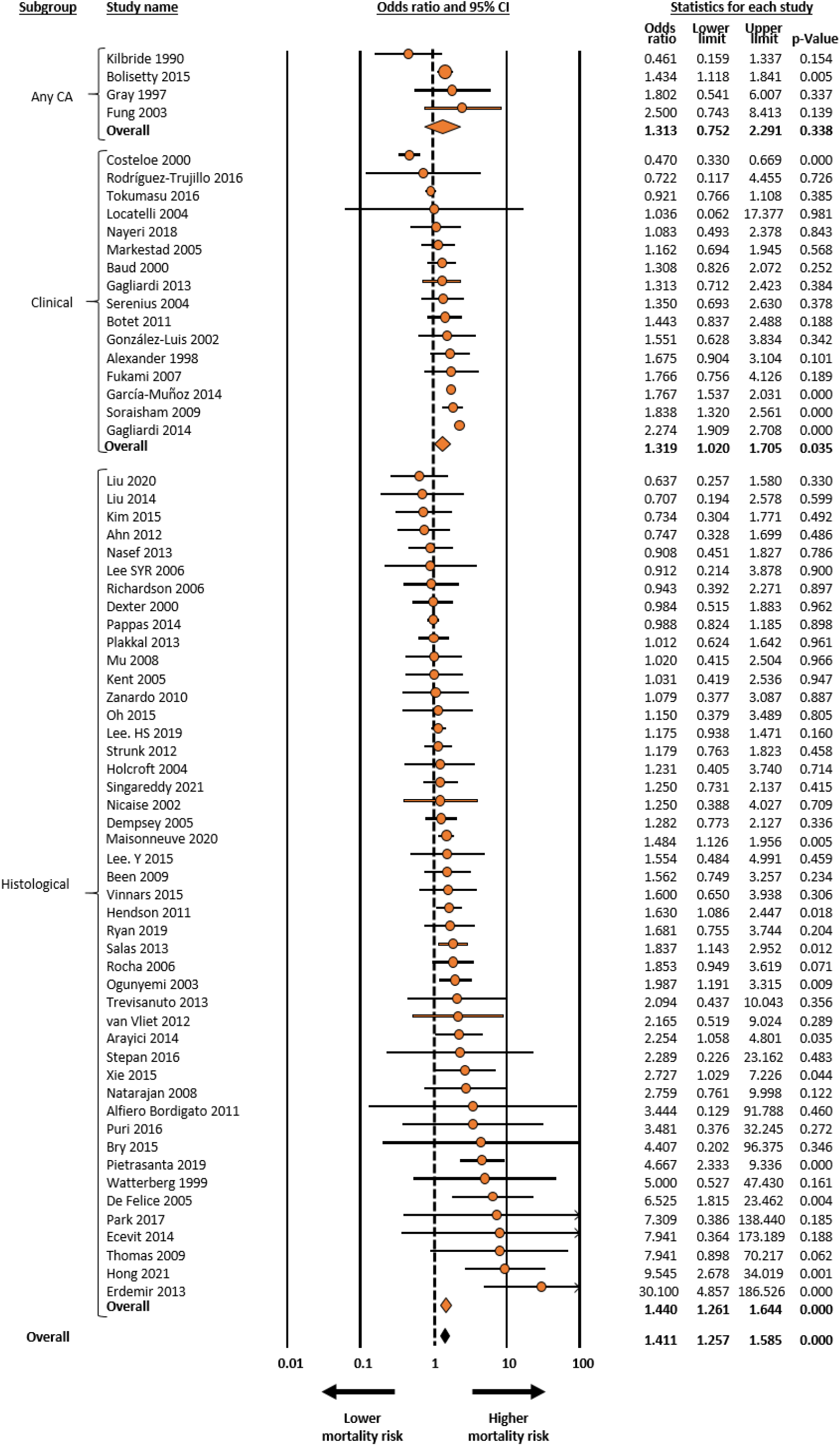
Meta-analysis on association between chorioamnionitis and mortality in infants with gestational age up to 34 weeks. CA: chorioamnionitis, CI: confidence interval.

**eFigure 3.**
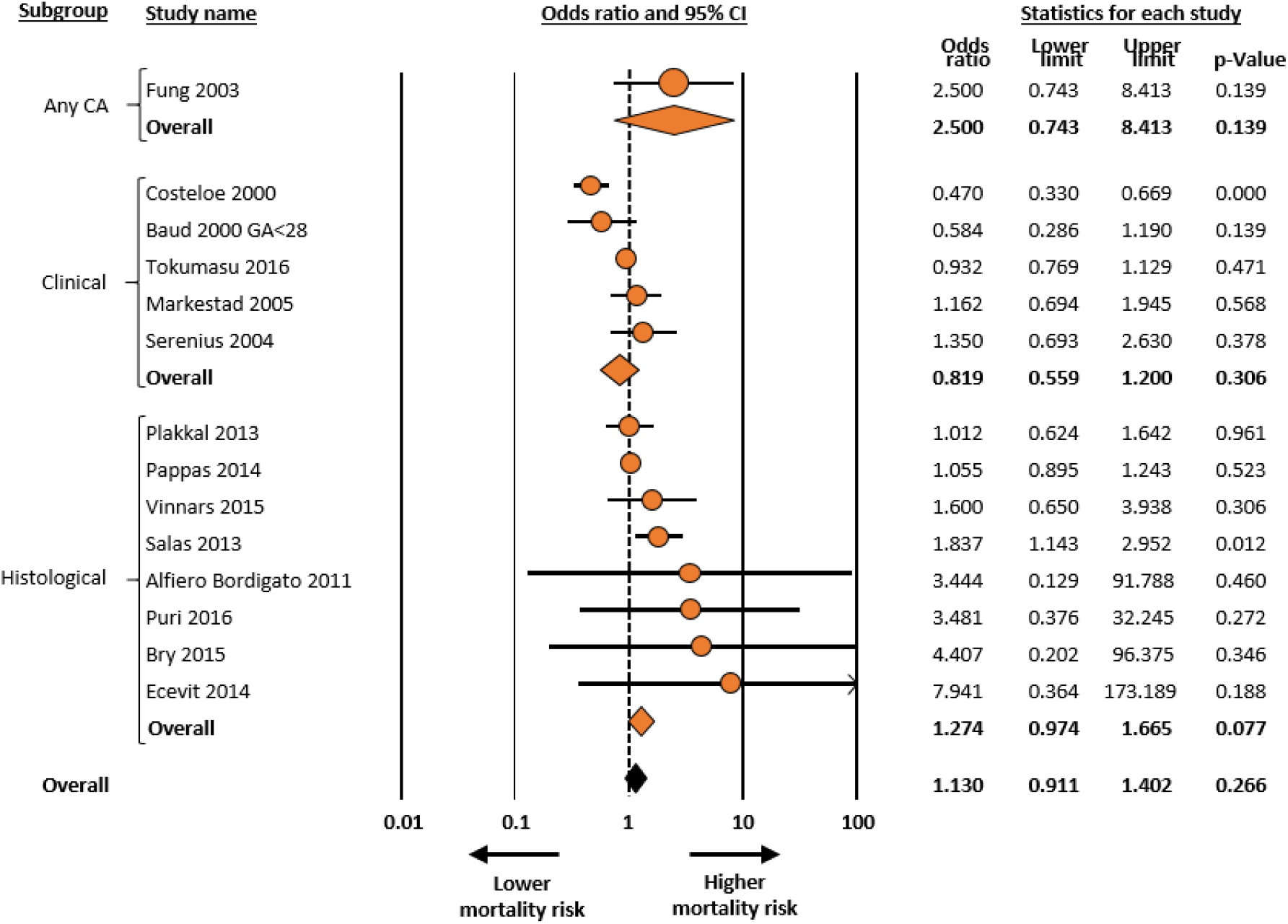
Meta-analysis on association between chorioamnionitis and mortality in infants with gestational age up to 28 weeks. CA: chorioamnionitis, CI: confidence interval

**eFigure 4.**
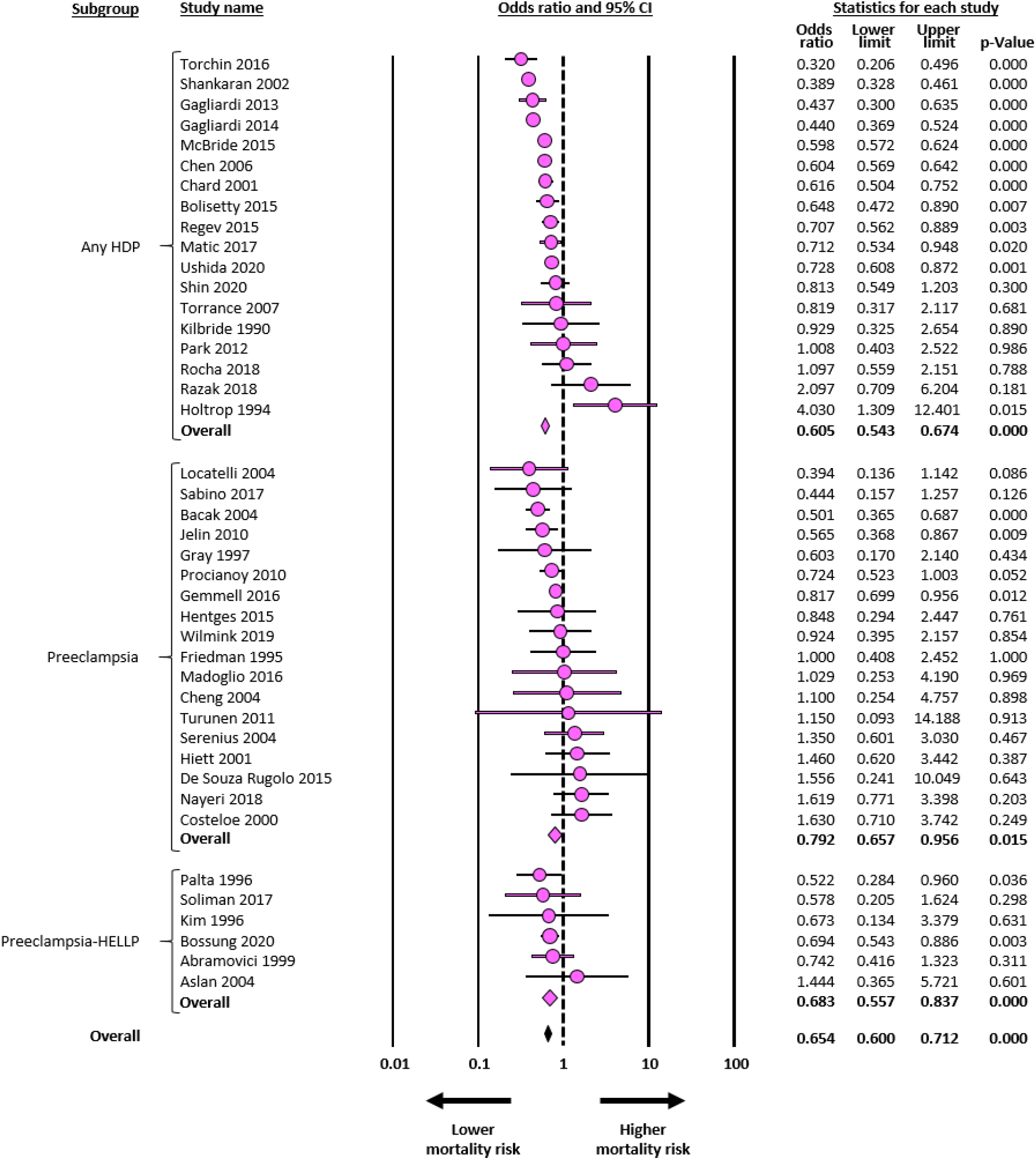
Meta-analysis on association between hypertensive disorders of pregnancy and mortality in infants with gestational age up to 34 weeks. HDP: hypertensive disorders of pregnancy, HELLP: syndrome in pregnancy characterized by hemolysis, elevated liver enzymes and low platelet count, CI: confidence interval.

**eFigure 5.**
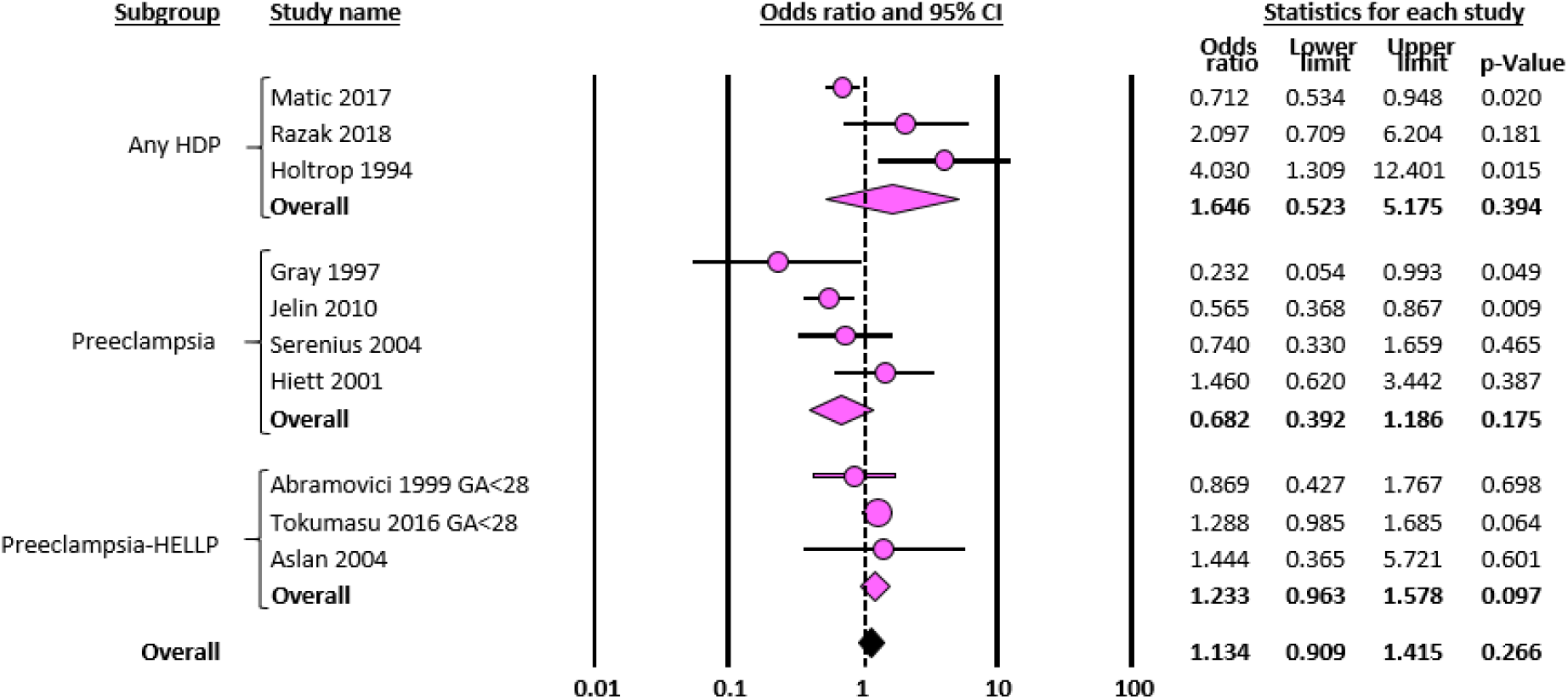
Meta-analysis on association between hypertensive disorders of pregnancy and mortality in infants with gestational age up to 28 weeks. HDP: hypertensive disorders of pregnancy, HELLP: syndrome in pregnancy characterized by hemolysis, elevated liver enzymes and low platelet count, CI: confidence interval.

**eFigure 6.**
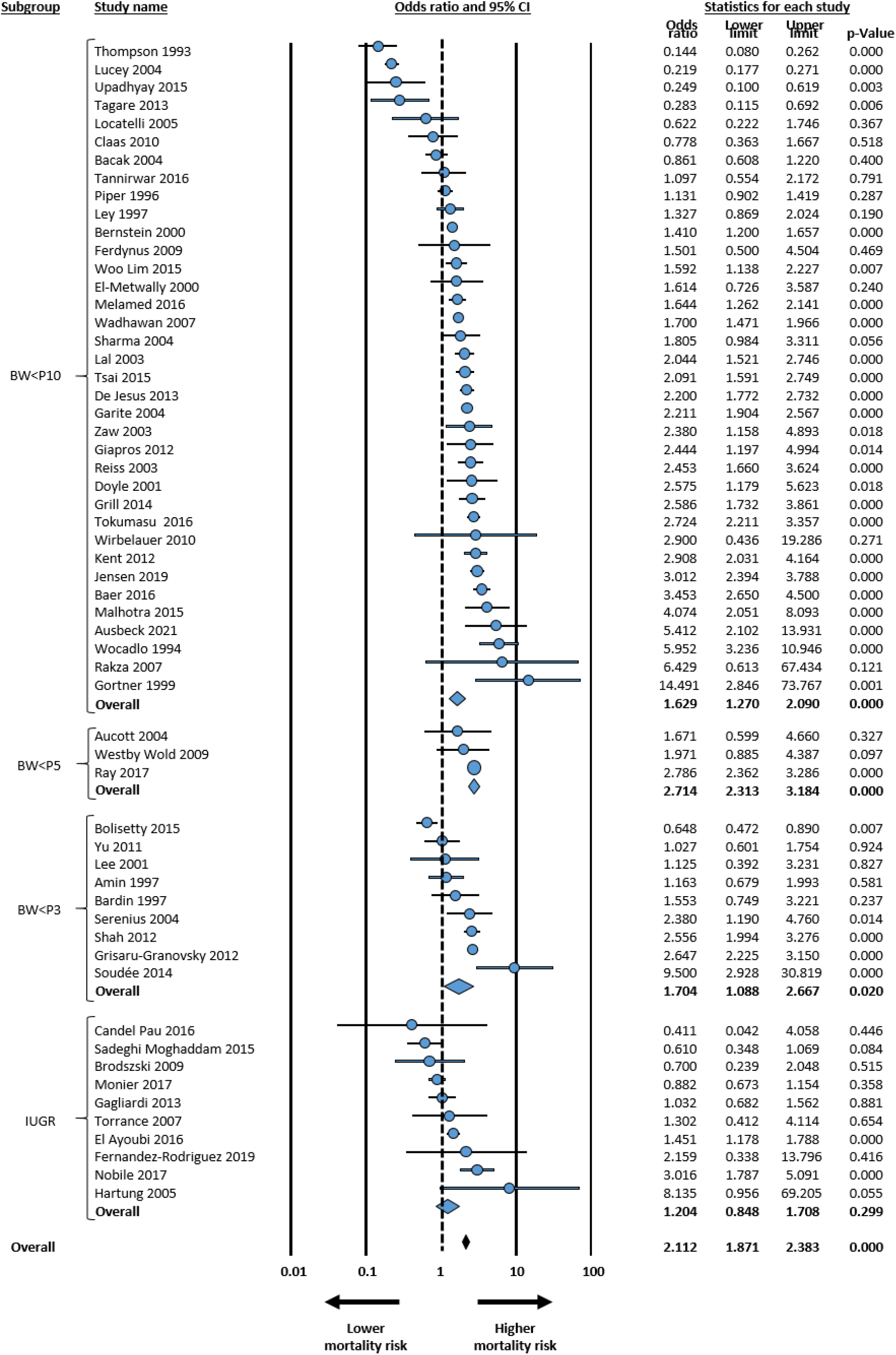
Meta-analysis on association between small for gestational age/intrauterine growth restriction and mortality in infants with gestational age up to 34 weeks. BW: birth weight, P10: 10^th^ percentile, P5: 5^th^ percentile, P3: 3^rd^ percentile; IUGR: intrauterine growth restriction defined on the basis of fetal growth assessment, CI: confidence interval.

**eFigure 7.**
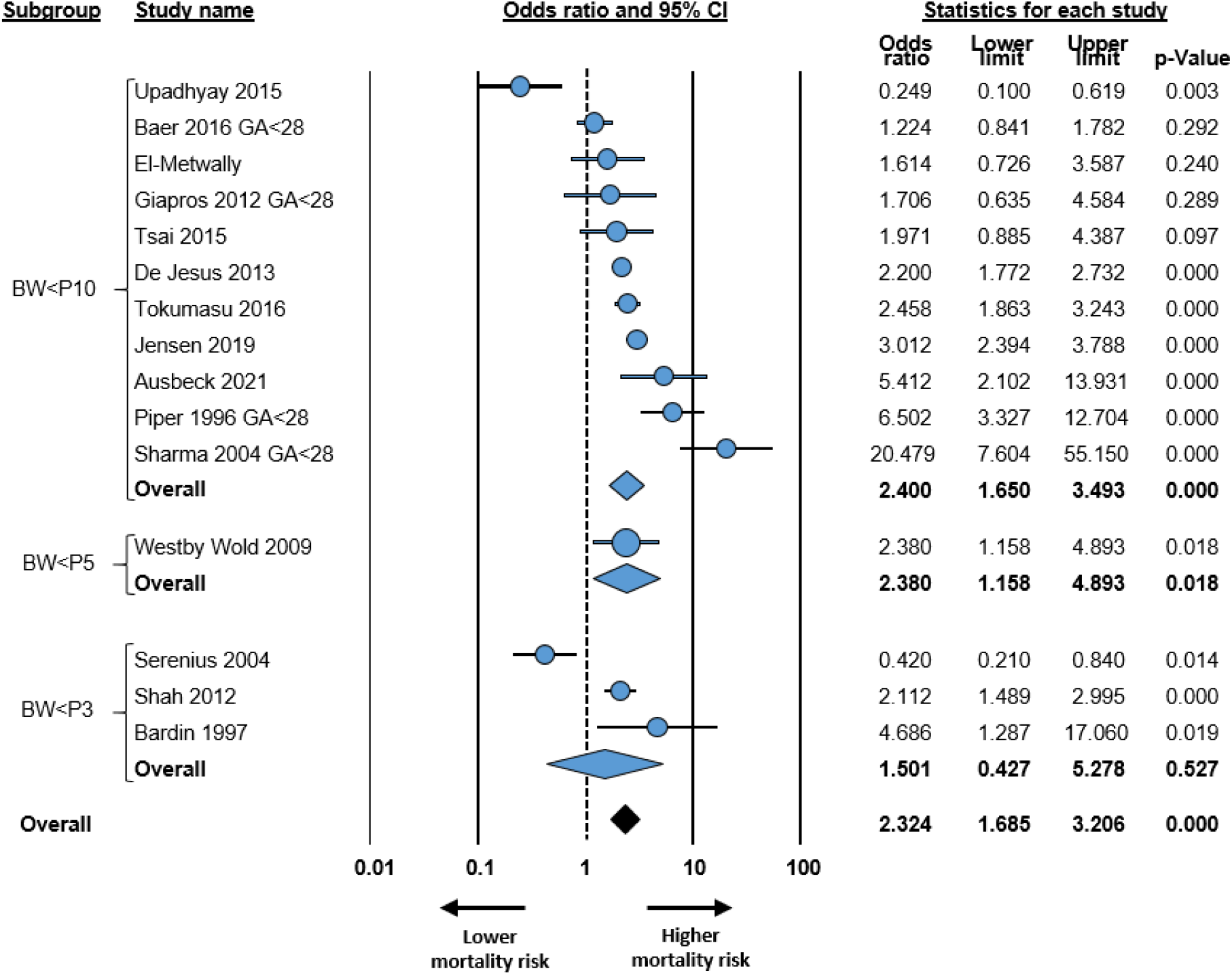
Meta-analysis on association between small for gestational age/intrauterine growth restriction and mortality in infants with gestational age up to 28 weeks. BW: birth weight, P10: 10^th^ percentile, P5: 5^th^ percentile, P3: 3^rd^ percentile, CI: confidence interval.

**eFigure 8.**
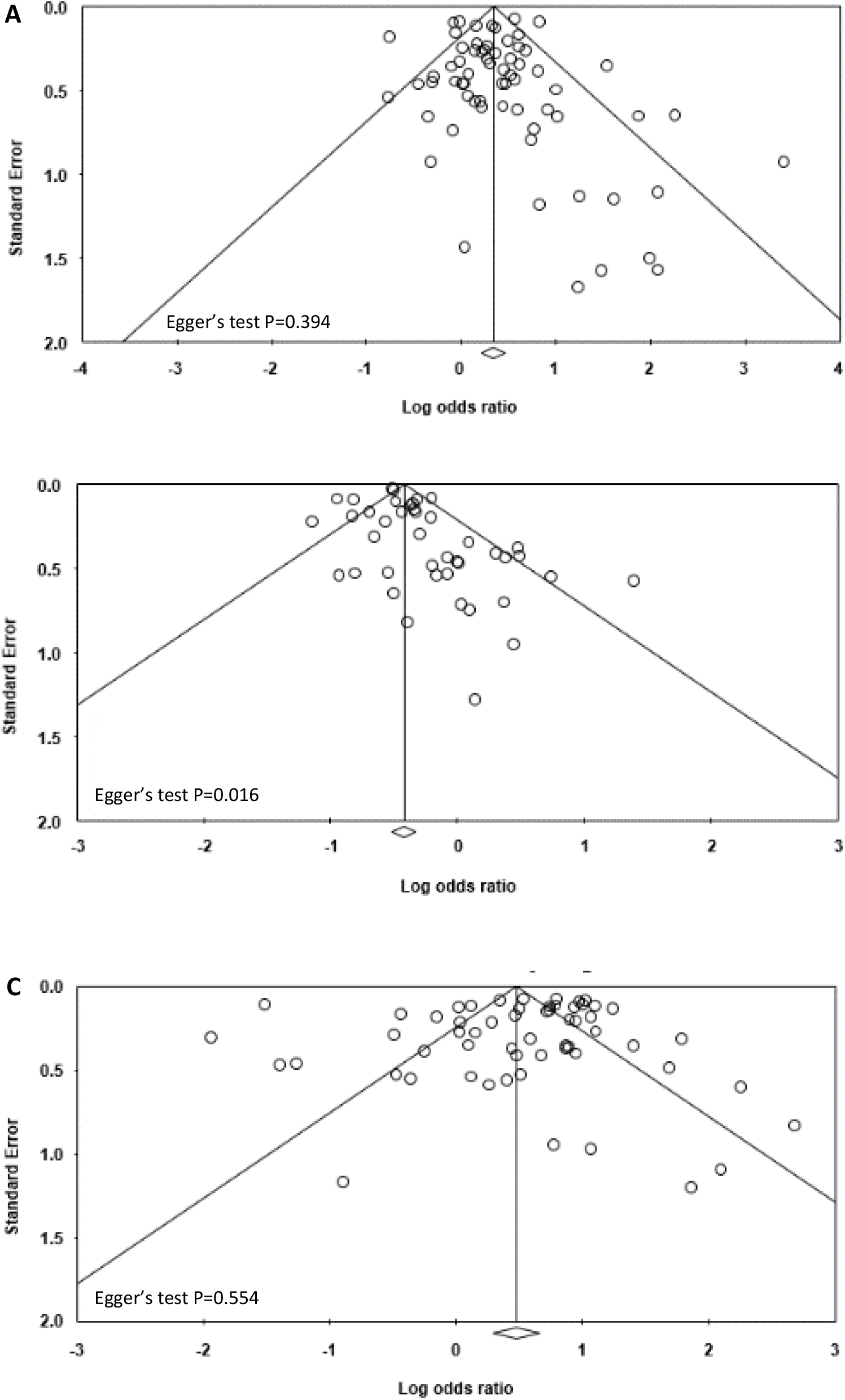
Funnel plot for publication bias analysis for the studies included in meta-analysis. **A**. Meta-analysis of chorioamnionitis and mortality **B**. Meta-analysis of hypertensive disorders of pregnancy (HDP) and mortality **C**. Meta-analysis of small for gestational age (SGA)/intrauterine growth restriction (IUGR) and mortality

**Table 1.**
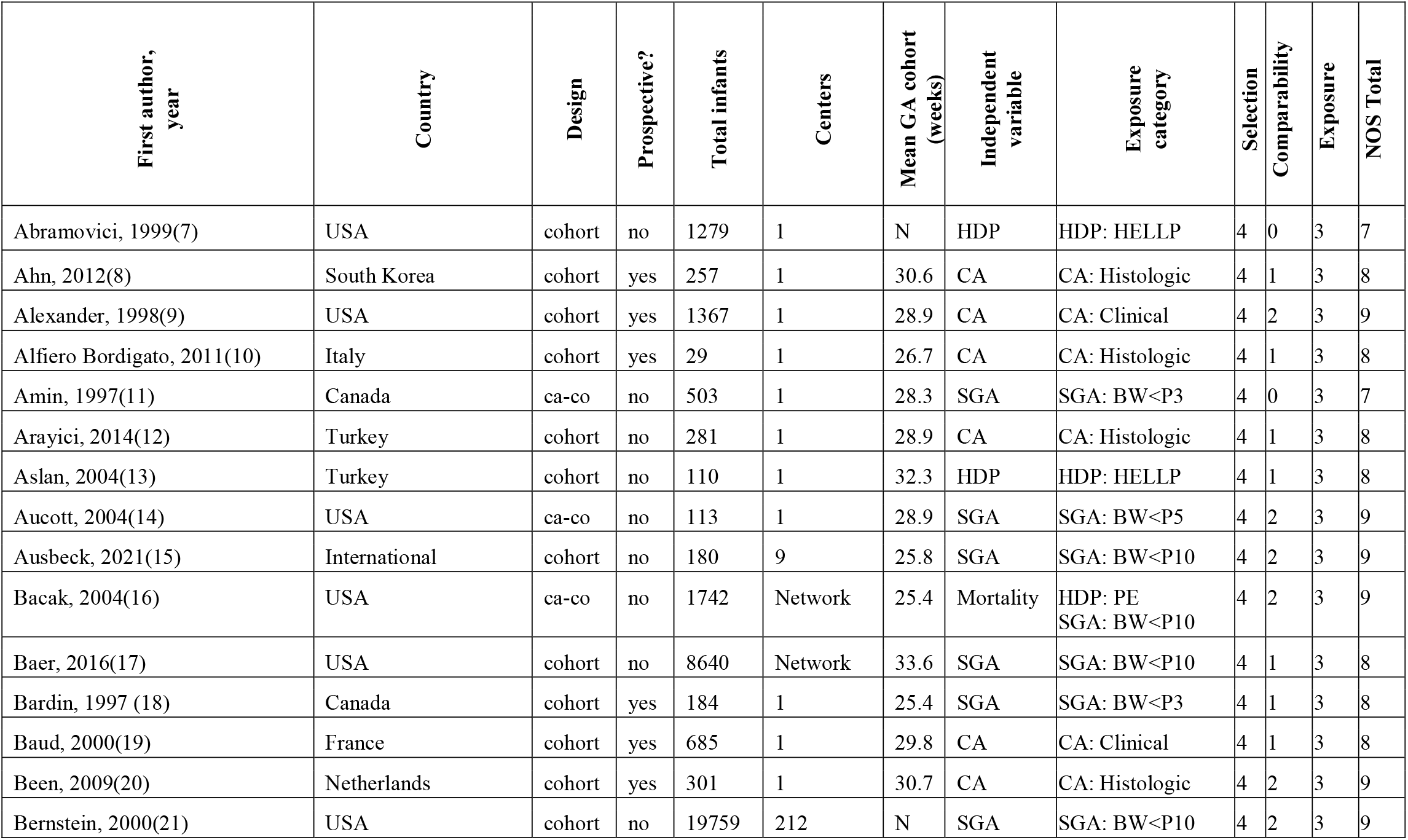
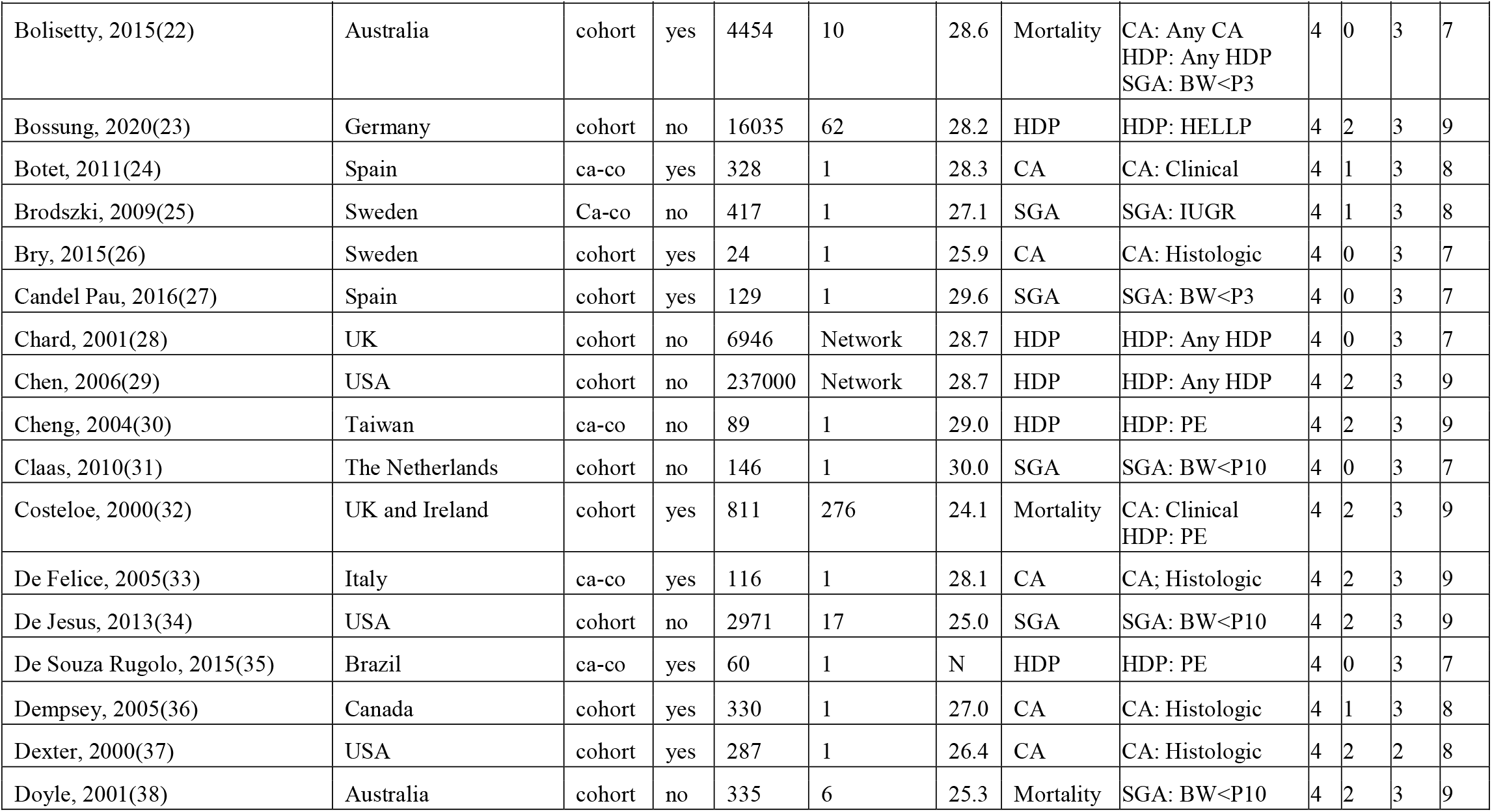
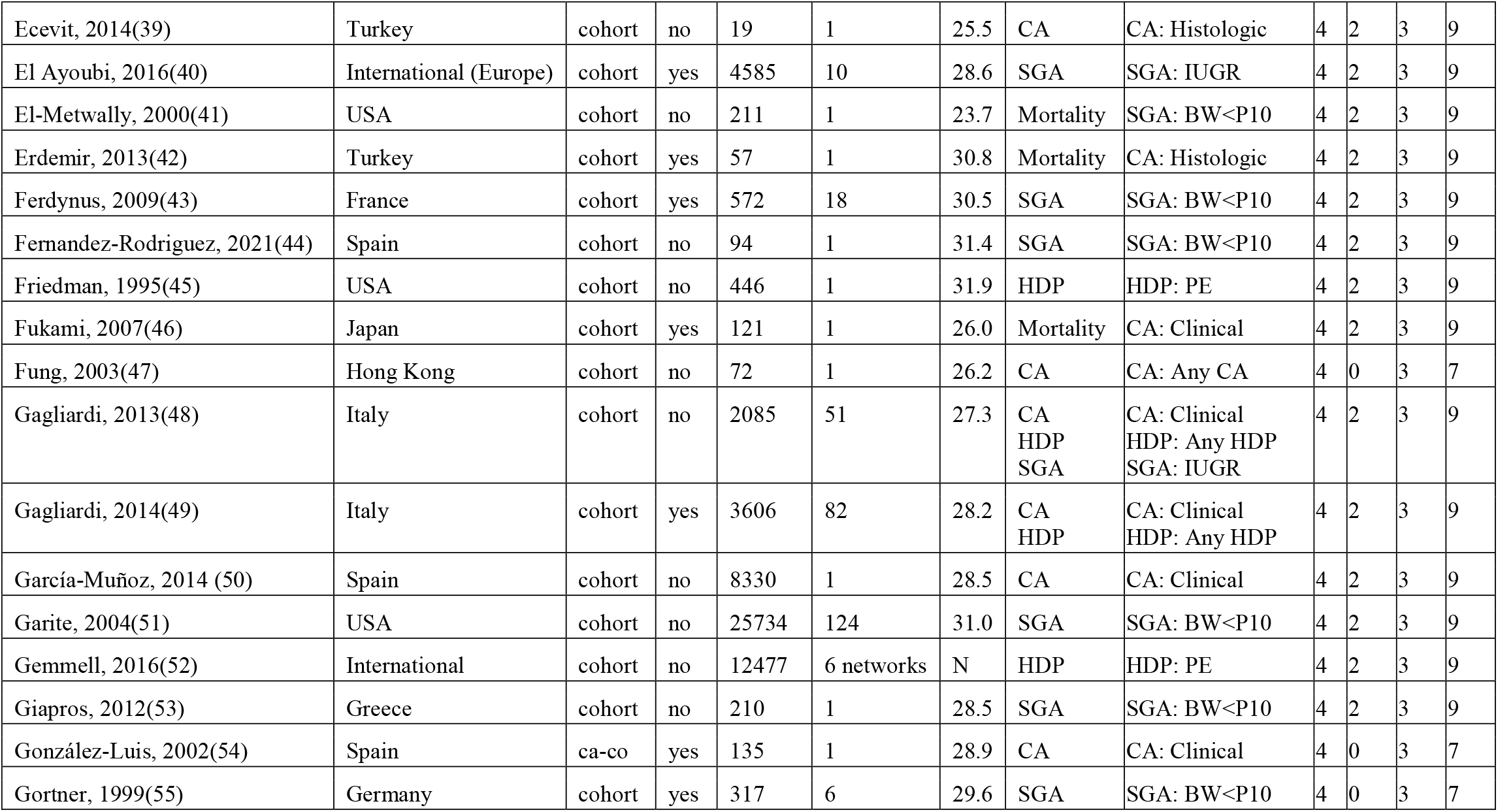
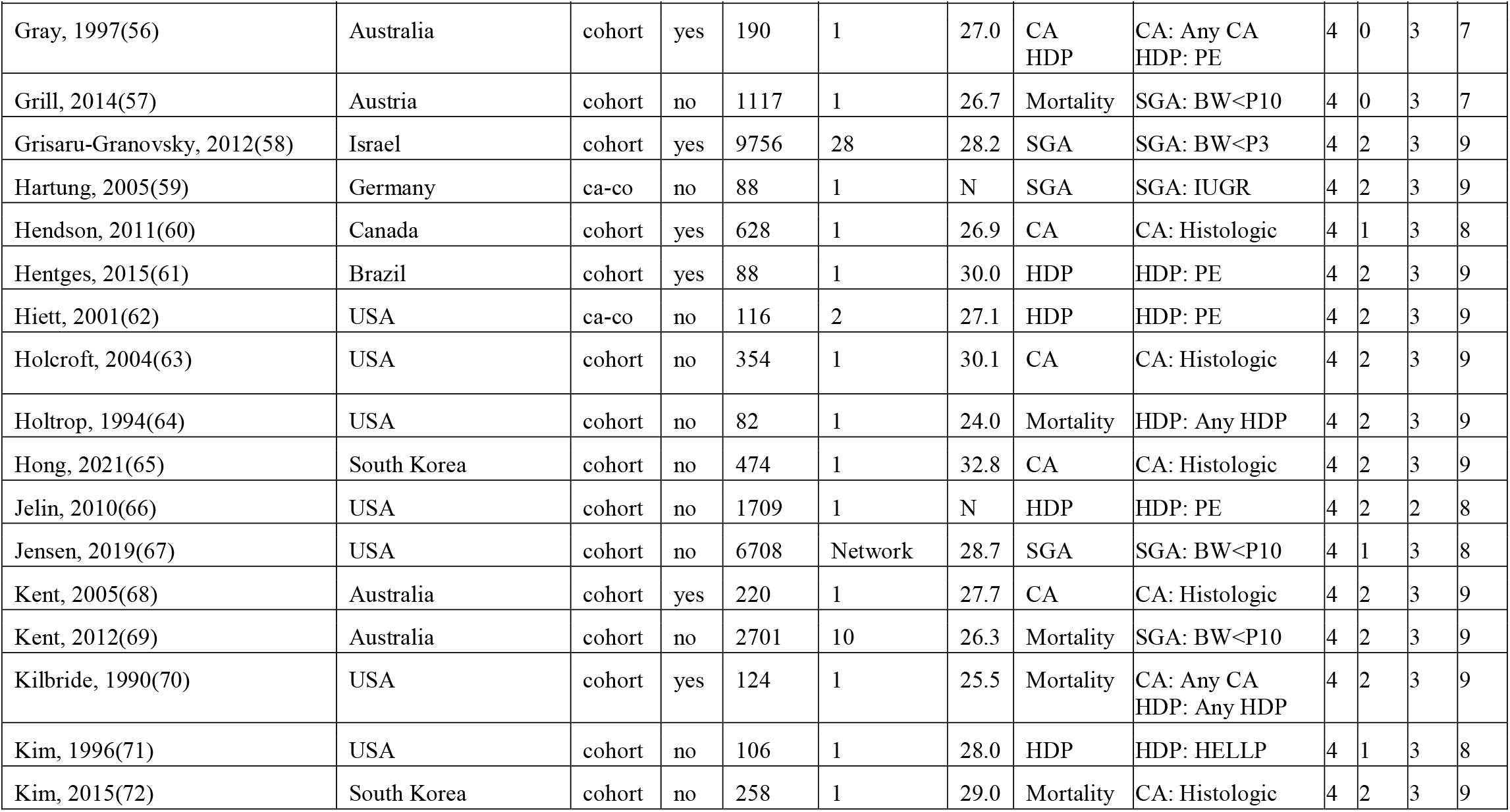
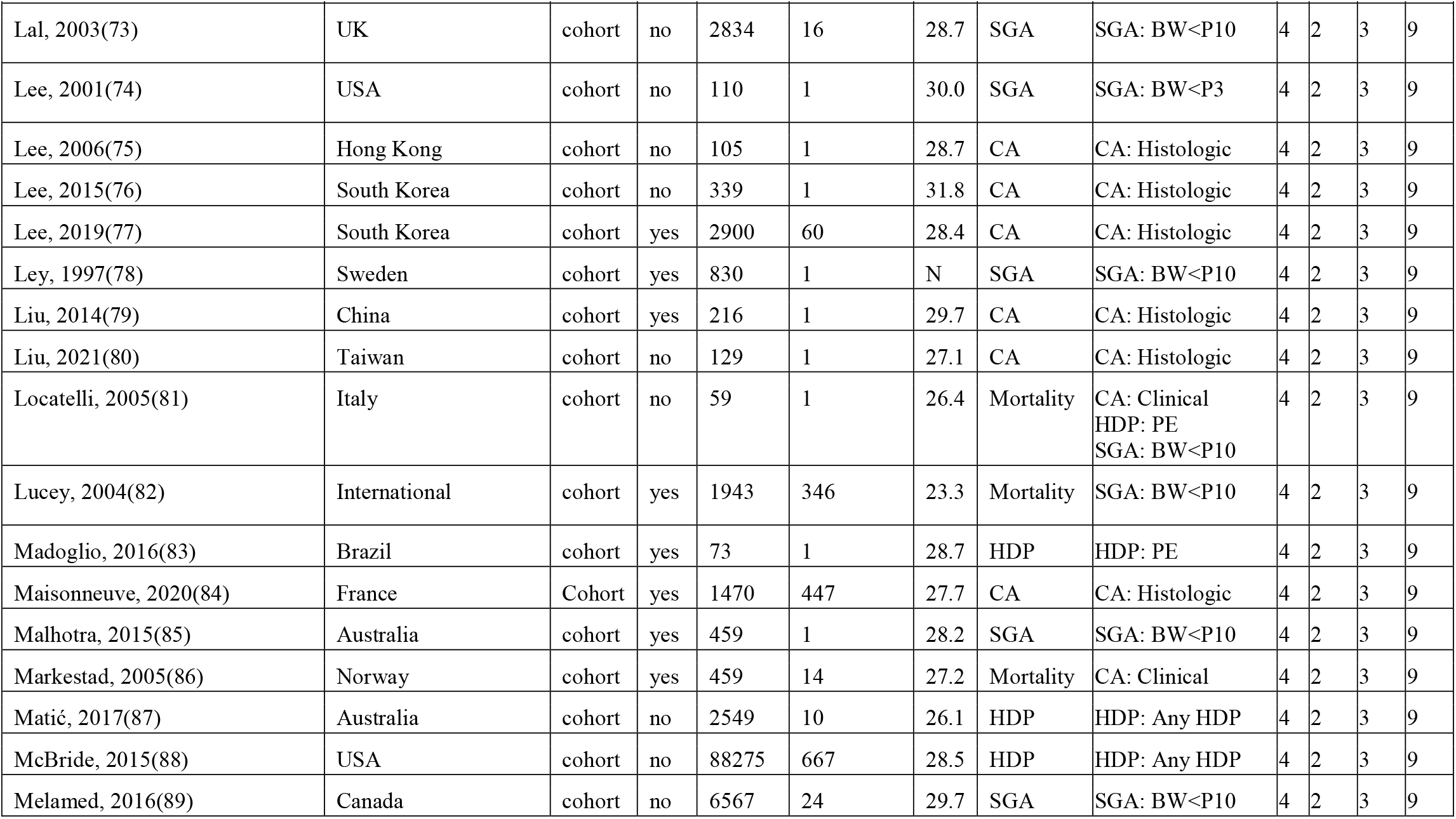
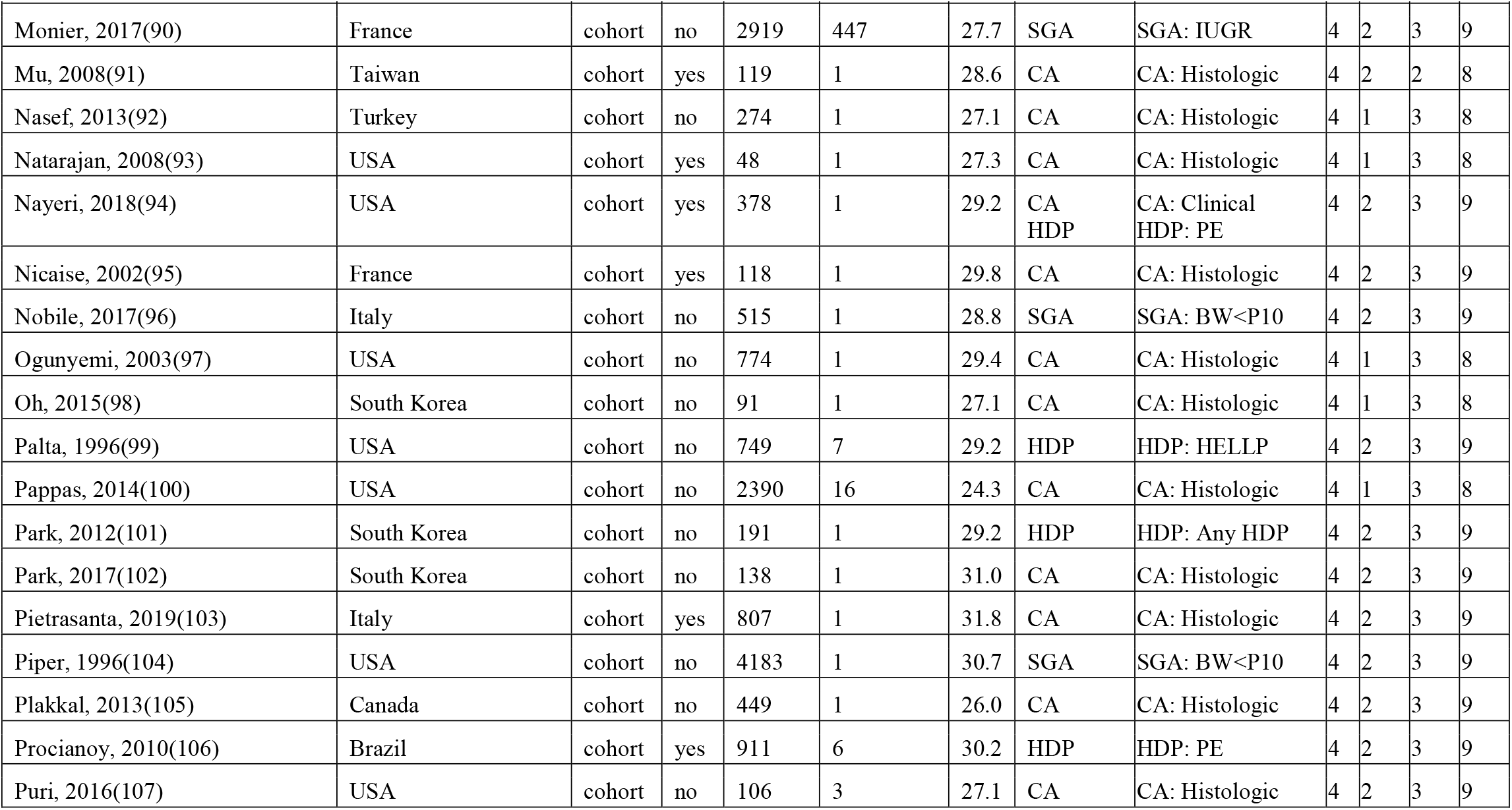
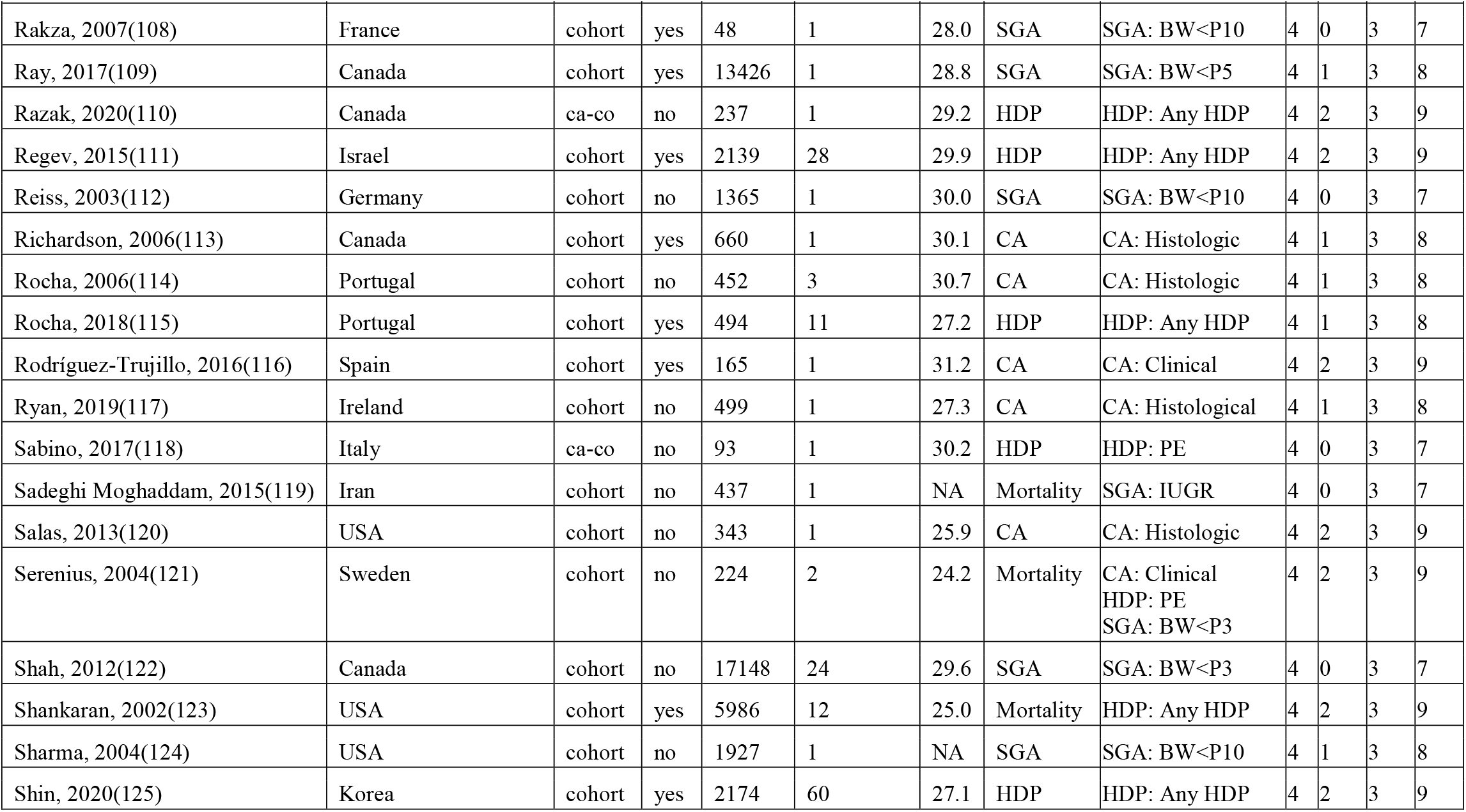
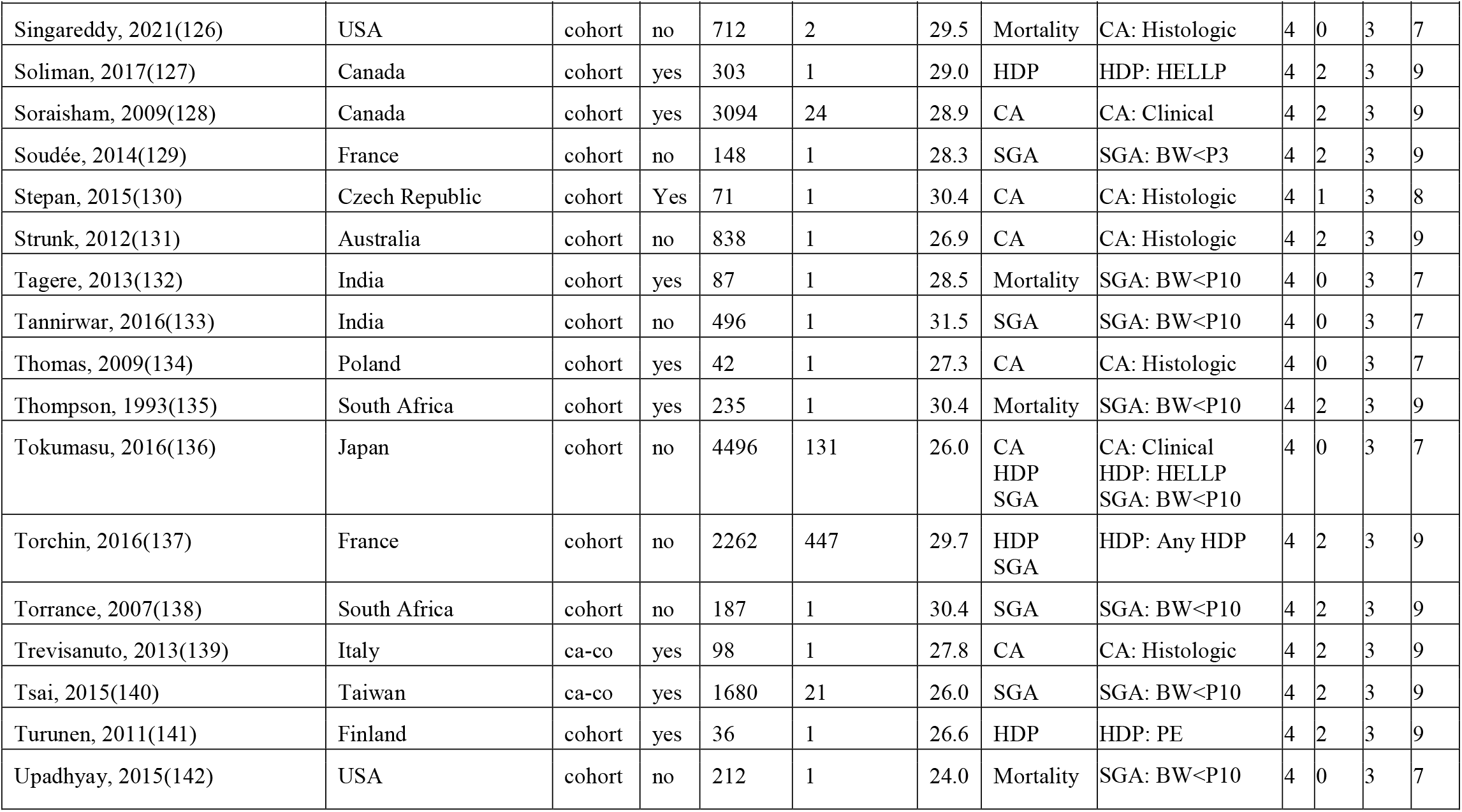
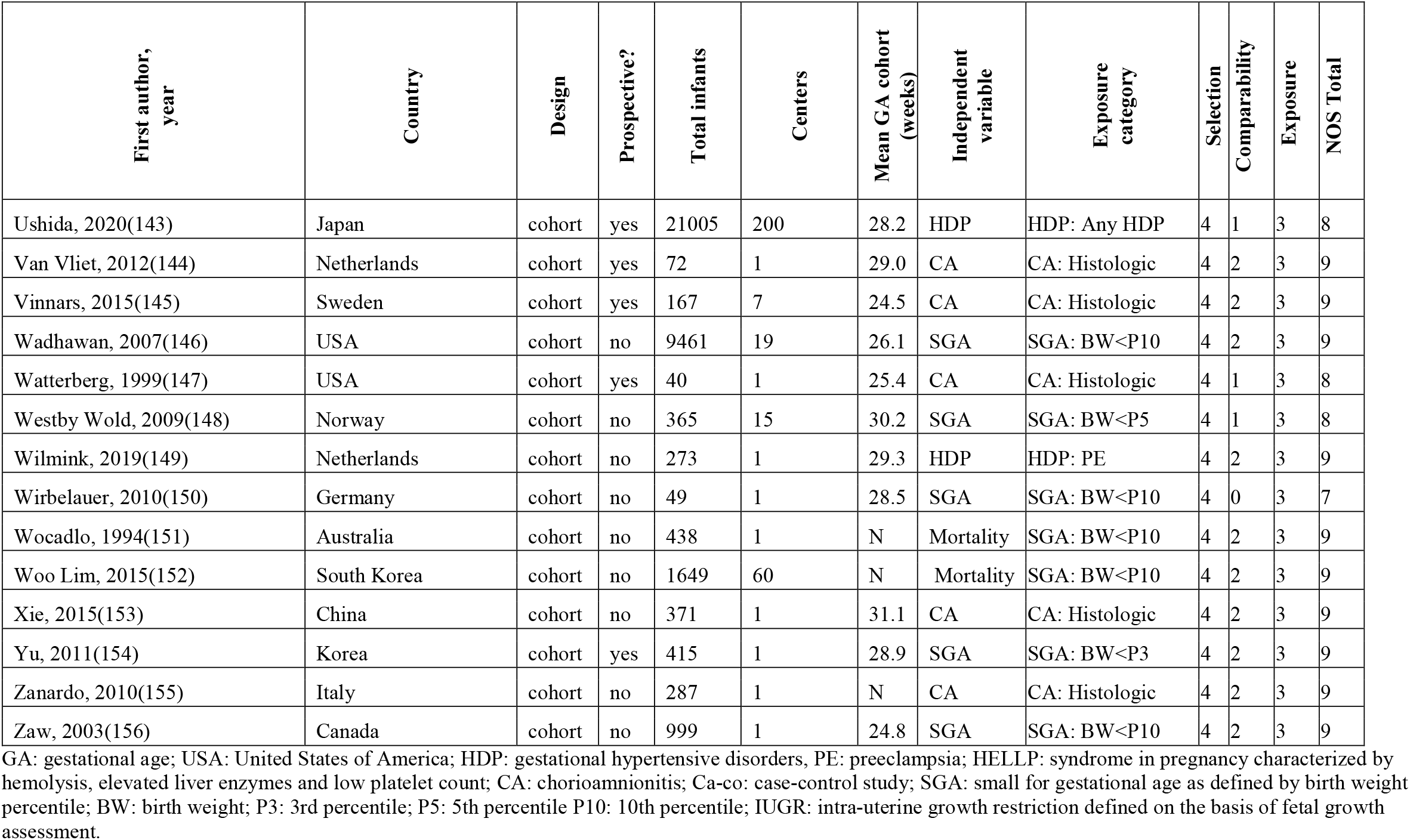
Characteristics of the included studies and Newcastle-Ottawa Scale.

| First author,<br>year | Country | Design | Prospective? | Total infants | Centers | Mean GA cohort<br>(weeks) | Independent<br>variable | Exposure<br>category | Selection | Comparability | Exposure | NOS Total |
| --- | --- | --- | --- | --- | --- | --- | --- | --- | --- | --- | --- | --- |
| Abramovici, 1999(7) | USA | cohort | no | 1279 | 1 | N | HDP | HDP: HELLP | 4 | 0 | 3 | 7 |
| Ahn, 2012(8) | South Korea | cohort | yes | 257 | 1 | 30.6 | CA | CA: Histologic | 4 | 1 | 3 | 8 |
| Alexander, 1998(9) | USA | cohort | yes | 1367 | 1 | 28.9 | CA | CA: Clinical | 4 | 2 | 3 | 9 |
| Alfiero Bordigato, 2011(10) | Italy | cohort | yes | 29 | 1 | 26.7 | CA | CA: Histologic | 4 | 1 | 3 | 8 |
| Amin, 1997(11) | Canada | ca-co | no | 503 | 1 | 28.3 | SGA | SGA: BW<P3 | 4 | 0 | 3 | 7 |
| Arayici, 2014(12) | Turkey | cohort | no | 281 | 1 | 28.9 | CA | CA: Histologic | 4 | 1 | 3 | 8 |
| Aslan, 2004(13) | Turkey | cohort | no | 110 | 1 | 32.3 | HDP | HDP: HELLP | 4 | 1 | 3 | 8 |
| Aucott, 2004(14) | USA | ca-co | no | 113 | 1 | 28.9 | SGA | SGA: BW<P5 | 4 | 2 | 3 | 9 |
| Ausbeck, 2021(15) | International | cohort | no | 180 | 9 | 25.8 | SGA | SGA: BW<P10 | 4 | 2 | 3 | 9 |
| Bacak, 2004(16) | USA | ca-co | no | 1742 | Network | 25.4 | Mortality | HDP: PE<br>SGA: BW<P10 | 4 | 2 | 3 | 9 |
| Baer, 2016(17) | USA | cohort | no | 8640 | Network | 33.6 | SGA | SGA: BW<P10 | 4 | 1 | 3 | 8 |
| Bardin, 1997 (18) | Canada | cohort | yes | 184 | 1 | 25.4 | SGA | SGA: BW<P3 | 4 | 1 | 3 | 8 |
| Baud, 2000(19) | France | cohort | yes | 685 | 1 | 29.8 | CA | CA: Clinical | 4 | 1 | 3 | 8 |
| Been, 2009(20) | Netherlands | cohort | yes | 301 | 1 | 30.7 | CA | CA: Histologic | 4 | 2 | 3 | 9 |
| Bernstein, 2000(21) | USA | cohort | no | 19759 | 212 | N | SGA | SGA: BW<P10 | 4 | 2 | 3 | 9 |
| Bolisetty, 2015(22) | Australia | cohort | yes | 4454 | 10 | 28.6 | Mortality | CA: Any CA<br>HDP: Any HDP<br>SGA: BW<P3 | 4 | 0 | 3 | 7 |
| Bossung, 2020(23) | Germany | cohort | no | 16035 | 62 | 28.2 | HDP | HDP: HELLP | 4 | 2 | 3 | 9 |
| Botet, 2011(24) | Spain | ca-co | yes | 328 | 1 | 28.3 | CA | CA: Clinical | 4 | 1 | 3 | 8 |
| Brodzski, 2009(25) | Sweden | Ca-co | no | 417 | 1 | 27.1 | SGA | SGA: IUGR | 4 | 1 | 3 | 8 |
| Bry, 2015(26) | Sweden | cohort | yes | 24 | 1 | 25.9 | CA | CA: Histologic | 4 | 0 | 3 | 7 |
| Candel Pau, 2016(27) | Spain | cohort | yes | 129 | 1 | 29.6 | SGA | SGA: BW<P3 | 4 | 0 | 3 | 7 |
| Chard, 2001(28) | UK | cohort | no | 6946 | Network | 28.7 | HDP | HDP: Any HDP | 4 | 0 | 3 | 7 |
| Chen, 2006(29) | USA | cohort | no | 237000 | Network | 28.7 | HDP | HDP: Any HDP | 4 | 2 | 3 | 9 |
| Cheng, 2004(30) | Taiwan | ca-co | no | 89 | 1 | 29.0 | HDP | HDP: PE | 4 | 2 | 3 | 9 |
| Claas, 2010(31) | The Netherlands | cohort | no | 146 | 1 | 30.0 | SGA | SGA: BW<P10 | 4 | 0 | 3 | 7 |
| Costeloe, 2000(32) | UK and Ireland | cohort | yes | 811 | 276 | 24.1 | Mortality | CA: Clinical<br>HDP: PE | 4 | 2 | 3 | 9 |
| De Felice, 2005(33) | Italy | ca-co | yes | 116 | 1 | 28.1 | CA | CA; Histologic | 4 | 2 | 3 | 9 |
| De Jesus, 2013(34) | USA | cohort | no | 2971 | 17 | 25.0 | SGA | SGA: BW<P10 | 4 | 2 | 3 | 9 |
| De Souza Rugolo, 2015(35) | Brazil | ca-co | yes | 60 | 1 | N | HDP | HDP: PE | 4 | 0 | 3 | 7 |
| Dempsey, 2005(36) | Canada | cohort | yes | 330 | 1 | 27.0 | CA | CA: Histologic | 4 | 1 | 3 | 8 |
| Dexter, 2000(37) | USA | cohort | yes | 287 | 1 | 26.4 | CA | CA: Histologic | 4 | 2 | 2 | 8 |
| Doyle, 2001(38) | Australia | cohort | no | 335 | 6 | 25.3 | Mortality | SGA: BW<P10 | 4 | 2 | 3 | 9 |
| Ecevit, 2014(39) | Turkey | cohort | no | 19 | 1 | 25.5 | CA | CA: Histologic | 4 | 2 | 3 | 9 |
| El Ayoubi, 2016(40) | International (Europe) | cohort | yes | 4585 | 10 | 28.6 | SGA | SGA: IUGR | 4 | 2 | 3 | 9 |
| El-Metwally, 2000(41) | USA | cohort | no | 211 | 1 | 23.7 | Mortality | SGA: BW<P10 | 4 | 2 | 3 | 9 |
| Erdemir, 2013(42) | Turkey | cohort | yes | 57 | 1 | 30.8 | Mortality | CA: Histologic | 4 | 2 | 3 | 9 |
| Ferdynus, 2009(43) | France | cohort | yes | 572 | 18 | 30.5 | SGA | SGA: BW<P10 | 4 | 2 | 3 | 9 |
| Fernandez-Rodriguez, 2021(44) | Spain | cohort | no | 94 | 1 | 31.4 | SGA | SGA: BW<P10 | 4 | 2 | 3 | 9 |
| Friedman, 1995(45) | USA | cohort | no | 446 | 1 | 31.9 | HDP | HDP: PE | 4 | 2 | 3 | 9 |
| Fukami, 2007(46) | Japan | cohort | yes | 121 | 1 | 26.0 | Mortality | CA: Clinical | 4 | 2 | 3 | 9 |
| Fung, 2003(47) | Hong Kong | cohort | no | 72 | 1 | 26.2 | CA | CA: Any CA | 4 | 0 | 3 | 7 |
| Gagliardi, 2013(48) | Italy | cohort | no | 2085 | 51 | 27.3 | CA<br>HDP<br>SGA | CA: Clinical<br>HDP: Any HDP<br>SGA: IUGR | 4 | 2 | 3 | 9 |
| Gagliardi, 2014(49) | Italy | cohort | yes | 3606 | 82 | 28.2 | CA<br>HDP | CA: Clinical<br>HDP: Any HDP | 4 | 2 | 3 | 9 |
| García-Muñoz, 2014 (50) | Spain | cohort | no | 8330 | 1 | 28.5 | CA | CA: Clinical | 4 | 2 | 3 | 9 |
| Garite, 2004(51) | USA | cohort | no | 25734 | 124 | 31.0 | SGA | SGA: BW<P10 | 4 | 2 | 3 | 9 |
| Gemmell, 2016(52) | International | cohort | no | 12477 | 6 networks | N | HDP | HDP: PE | 4 | 2 | 3 | 9 |
| Giapros, 2012(53) | Greece | cohort | no | 210 | 1 | 28.5 | SGA | SGA: BW<P10 | 4 | 2 | 3 | 9 |
| González-Luis, 2002(54) | Spain | ca-co | yes | 135 | 1 | 28.9 | CA | CA: Clinical | 4 | 0 | 3 | 7 |
| Gortner, 1999(55) | Germany | cohort | yes | 317 | 6 | 29.6 | SGA | SGA: BW<P10 | 4 | 0 | 3 | 7 |
| Gray, 1997(56) | Australia | cohort | yes | 190 | 1 | 27.0 | CA<br>HDP | CA: Any CA<br>HDP: PE | 4 | 0 | 3 | 7 |
| Grill, 2014(57) | Austria | cohort | no | 1117 | 1 | 26.7 | Mortality | SGA: BW<P10 | 4 | 0 | 3 | 7 |
| Grisaru-Granovsky, 2012(58) | Israel | cohort | yes | 9756 | 28 | 28.2 | SGA | SGA: BW<P3 | 4 | 2 | 3 | 9 |
| Hartung, 2005(59) | Germany | ca-co | no | 88 | 1 | N | SGA | SGA: IUGR | 4 | 2 | 3 | 9 |
| Hendson, 2011(60) | Canada | cohort | yes | 628 | 1 | 26.9 | CA | CA: Histologic | 4 | 1 | 3 | 8 |
| Hentges, 2015(61) | Brazil | cohort | yes | 88 | 1 | 30.0 | HDP | HDP: PE | 4 | 2 | 3 | 9 |
| Hiett, 2001(62) | USA | ca-co | no | 116 | 2 | 27.1 | HDP | HDP: PE | 4 | 2 | 3 | 9 |
| Holcroft, 2004(63) | USA | cohort | no | 354 | 1 | 30.1 | CA | CA: Histologic | 4 | 2 | 3 | 9 |
| Holtrop, 1994(64) | USA | cohort | no | 82 | 1 | 24.0 | Mortality | HDP: Any HDP | 4 | 2 | 3 | 9 |
| Hong, 2021(65) | South Korea | cohort | no | 474 | 1 | 32.8 | CA | CA: Histologic | 4 | 2 | 3 | 9 |
| Jelin, 2010(66) | USA | cohort | no | 1709 | 1 | N | HDP | HDP: PE | 4 | 2 | 2 | 8 |
| Jensen, 2019(67) | USA | cohort | no | 6708 | Network | 28.7 | SGA | SGA: BW<P10 | 4 | 1 | 3 | 8 |
| Kent, 2005(68) | Australia | cohort | yes | 220 | 1 | 27.7 | CA | CA: Histologic | 4 | 2 | 3 | 9 |
| Kent, 2012(69) | Australia | cohort | no | 2701 | 10 | 26.3 | Mortality | SGA: BW<P10 | 4 | 2 | 3 | 9 |
| Kilbride, 1990(70) | USA | cohort | yes | 124 | 1 | 25.5 | Mortality | CA: Any CA<br>HDP: Any HDP | 4 | 2 | 3 | 9 |
| Kim, 1996(71) | USA | cohort | no | 106 | 1 | 28.0 | HDP | HDP: HELLP | 4 | 1 | 3 | 8 |
| Kim, 2015(72) | South Korea | cohort | no | 258 | 1 | 29.0 | Mortality | CA: Histologic | 4 | 2 | 3 | 9 |
| Lal, 2003(73) | UK | cohort | no | 2834 | 16 | 28.7 | SGA | SGA: BW<P10 | 4 | 2 | 3 | 9 |
| Lee, 2001(74) | USA | cohort | no | 110 | 1 | 30.0 | SGA | SGA: BW<P3 | 4 | 2 | 3 | 9 |
| Lee, 2006(75) | Hong Kong | cohort | no | 105 | 1 | 28.7 | CA | CA: Histologic | 4 | 2 | 3 | 9 |
| Lee, 2015(76) | South Korea | cohort | no | 339 | 1 | 31.8 | CA | CA: Histologic | 4 | 2 | 3 | 9 |
| Lee, 2019(77) | South Korea | cohort | yes | 2900 | 60 | 28.4 | CA | CA: Histologic | 4 | 2 | 3 | 9 |
| Ley, 1997(78) | Sweden | cohort | yes | 830 | 1 | N | SGA | SGA: BW<P10 | 4 | 2 | 3 | 9 |
| Liu, 2014(79) | China | cohort | yes | 216 | 1 | 29.7 | CA | CA: Histologic | 4 | 2 | 3 | 9 |
| Liu, 2021(80) | Taiwan | cohort | no | 129 | 1 | 27.1 | CA | CA: Histologic | 4 | 2 | 3 | 9 |
| Locatelli, 2005(81) | Italy | cohort | no | 59 | 1 | 26.4 | Mortality | CA: Clinical<br>HDP: PE<br>SGA: BW<P10 | 4 | 2 | 3 | 9 |
| Lucey, 2004(82) | International | cohort | yes | 1943 | 346 | 23.3 | Mortality | SGA: BW<P10 | 4 | 2 | 3 | 9 |
| Madoglio, 2016(83) | Brazil | cohort | yes | 73 | 1 | 28.7 | HDP | HDP: PE | 4 | 2 | 3 | 9 |
| Maisonneuve, 2020(84) | France | Cohort | yes | 1470 | 447 | 27.7 | CA | CA: Histologic | 4 | 2 | 3 | 9 |
| Malhotra, 2015(85) | Australia | cohort | yes | 459 | 1 | 28.2 | SGA | SGA: BW<P10 | 4 | 2 | 3 | 9 |
| Markestad, 2005(86) | Norway | cohort | yes | 459 | 14 | 27.2 | Mortality | CA: Clinical | 4 | 2 | 3 | 9 |
| Matić, 2017(87) | Australia | cohort | no | 2549 | 10 | 26.1 | HDP | HDP: Any HDP | 4 | 2 | 3 | 9 |
| McBride, 2015(88) | USA | cohort | no | 88275 | 667 | 28.5 | HDP | HDP: Any HDP | 4 | 2 | 3 | 9 |
| Melamed, 2016(89) | Canada | cohort | no | 6567 | 24 | 29.7 | SGA | SGA: BW<P10 | 4 | 2 | 3 | 9 |
| Monier, 2017(90) | France | cohort | no | 2919 | 447 | 27.7 | SGA | SGA: IUGR | 4 | 2 | 3 | 9 |
| Mu, 2008(91) | Taiwan | cohort | yes | 119 | 1 | 28.6 | CA | CA: Histologic | 4 | 2 | 2 | 8 |
| Nasef, 2013(92) | Turkey | cohort | no | 274 | 1 | 27.1 | CA | CA: Histologic | 4 | 1 | 3 | 8 |
| Natarajan, 2008(93) | USA | cohort | yes | 48 | 1 | 27.3 | CA | CA: Histologic | 4 | 1 | 3 | 8 |
| Nayeri, 2018(94) | USA | cohort | yes | 378 | 1 | 29.2 | CA<br>HDP | CA: Clinical<br>HDP: PE | 4 | 2 | 3 | 9 |
| Nicaise, 2002(95) | France | cohort | yes | 118 | 1 | 29.8 | CA | CA: Histologic | 4 | 2 | 3 | 9 |
| Nobile, 2017(96) | Italy | cohort | no | 515 | 1 | 28.8 | SGA | SGA: BW<P10 | 4 | 2 | 3 | 9 |
| Ogunyemi, 2003(97) | USA | cohort | no | 774 | 1 | 29.4 | CA | CA: Histologic | 4 | 1 | 3 | 8 |
| Oh, 2015(98) | South Korea | cohort | no | 91 | 1 | 27.1 | CA | CA: Histologic | 4 | 1 | 3 | 8 |
| Palta, 1996(99) | USA | cohort | no | 749 | 7 | 29.2 | HDP | HDP: HELLP | 4 | 2 | 3 | 9 |
| Pappas, 2014(100) | USA | cohort | no | 2390 | 16 | 24.3 | CA | CA: Histologic | 4 | 1 | 3 | 8 |
| Park, 2012(101) | South Korea | cohort | no | 191 | 1 | 29.2 | HDP | HDP: Any HDP | 4 | 2 | 3 | 9 |
| Park, 2017(102) | South Korea | cohort | no | 138 | 1 | 31.0 | CA | CA: Histologic | 4 | 2 | 3 | 9 |
| Pietrasanta, 2019(103) | Italy | cohort | yes | 807 | 1 | 31.8 | CA | CA: Histologic | 4 | 2 | 3 | 9 |
| Piper, 1996(104) | USA | cohort | no | 4183 | 1 | 30.7 | SGA | SGA: BW<P10 | 4 | 2 | 3 | 9 |
| Plakkal, 2013(105) | Canada | cohort | no | 449 | 1 | 26.0 | CA | CA: Histologic | 4 | 2 | 3 | 9 |
| Procianoy, 2010(106) | Brazil | cohort | yes | 911 | 6 | 30.2 | HDP | HDP: PE | 4 | 2 | 3 | 9 |
| Puri, 2016(107) | USA | cohort | no | 106 | 3 | 27.1 | CA | CA: Histologic | 4 | 2 | 3 | 9 |
| Rakza, 2007(108) | France | cohort | yes | 48 | 1 | 28.0 | SGA | SGA: BW<P10 | 4 | 0 | 3 | 7 |
| Ray, 2017(109) | Canada | cohort | yes | 13426 | 1 | 28.8 | SGA | SGA: BW<P5 | 4 | 1 | 3 | 8 |
| Razak, 2020(110) | Canada | ca-co | no | 237 | 1 | 29.2 | HDP | HDP: Any HDP | 4 | 2 | 3 | 9 |
| Regev, 2015(111) | Israel | cohort | yes | 2139 | 28 | 29.9 | HDP | HDP: Any HDP | 4 | 2 | 3 | 9 |
| Reiss, 2003(112) | Germany | cohort | no | 1365 | 1 | 30.0 | SGA | SGA: BW<P10 | 4 | 0 | 3 | 7 |
| Richardson, 2006(113) | Canada | cohort | yes | 660 | 1 | 30.1 | CA | CA: Histologic | 4 | 1 | 3 | 8 |
| Rocha, 2006(114) | Portugal | cohort | no | 452 | 3 | 30.7 | CA | CA: Histologic | 4 | 1 | 3 | 8 |
| Rocha, 2018(115) | Portugal | cohort | yes | 494 | 11 | 27.2 | HDP | HDP: Any HDP | 4 | 1 | 3 | 8 |
| Rodríguez-Trujillo, 2016(116) | Spain | cohort | yes | 165 | 1 | 31.2 | CA | CA: Clinical | 4 | 2 | 3 | 9 |
| Ryan, 2019(117) | Ireland | cohort | no | 499 | 1 | 27.3 | CA | CA: Histological | 4 | 1 | 3 | 8 |
| Sabino, 2017(118) | Italy | ca-co | no | 93 | 1 | 30.2 | HDP | HDP: PE | 4 | 0 | 3 | 7 |
| Sadeghi Moghaddam, 2015(119) | Iran | cohort | no | 437 | 1 | NA | Mortality | SGA: IUGR | 4 | 0 | 3 | 7 |
| Salas, 2013(120) | USA | cohort | no | 343 | 1 | 25.9 | CA | CA: Histologic | 4 | 2 | 3 | 9 |
| Serenius, 2004(121) | Sweden | cohort | no | 224 | 2 | 24.2 | Mortality | CA: Clinical<br>HDP: PE<br>SGA: BW<P3 | 4 | 2 | 3 | 9 |
| Shah, 2012(122) | Canada | cohort | no | 17148 | 24 | 29.6 | SGA | SGA: BW<P3 | 4 | 0 | 3 | 7 |
| Shankaran, 2002(123) | USA | cohort | yes | 5986 | 12 | 25.0 | Mortality | HDP: Any HDP | 4 | 2 | 3 | 9 |
| Sharma, 2004(124) | USA | cohort | no | 1927 | 1 | NA | SGA | SGA: BW<P10 | 4 | 1 | 3 | 8 |
| Shin, 2020(125) | Korea | cohort | yes | 2174 | 60 | 27.1 | HDP | HDP: Any HDP | 4 | 2 | 3 | 9 |
| Singareddy, 2021(126) | USA | cohort | no | 712 | 2 | 29.5 | Mortality | CA: Histologic | 4 | 0 | 3 | 7 |
| Soliman, 2017(127) | Canada | cohort | yes | 303 | 1 | 29.0 | HDP | HDP: HELLP | 4 | 2 | 3 | 9 |
| Soraisham, 2009(128) | Canada | cohort | yes | 3094 | 24 | 28.9 | CA | CA: Clinical | 4 | 2 | 3 | 9 |
| Soudée, 2014(129) | France | cohort | no | 148 | 1 | 28.3 | SGA | SGA: BW<P3 | 4 | 2 | 3 | 9 |
| Stepan, 2015(130) | Czech Republic | cohort | Yes | 71 | 1 | 30.4 | CA | CA: Histologic | 4 | 1 | 3 | 8 |
| Strunk, 2012(131) | Australia | cohort | no | 838 | 1 | 26.9 | CA | CA: Histologic | 4 | 2 | 3 | 9 |
| Tagere, 2013(132) | India | cohort | yes | 87 | 1 | 28.5 | Mortality | SGA: BW<P10 | 4 | 0 | 3 | 7 |
| Tannirwar, 2016(133) | India | cohort | no | 496 | 1 | 31.5 | SGA | SGA: BW<P10 | 4 | 0 | 3 | 7 |
| Thomas, 2009(134) | Poland | cohort | yes | 42 | 1 | 27.3 | CA | CA: Histologic | 4 | 0 | 3 | 7 |
| Thompson, 1993(135) | South Africa | cohort | yes | 235 | 1 | 30.4 | Mortality | SGA: BW<P10 | 4 | 2 | 3 | 9 |
| Tokumasu, 2016(136) | Japan | cohort | no | 4496 | 131 | 26.0 | CA<br>HDP<br>SGA | CA: Clinical<br>HDP: HELLP<br>SGA: BW<P10 | 4 | 0 | 3 | 7 |
| Torchin, 2016(137) | France | cohort | no | 2262 | 447 | 29.7 | HDP<br>SGA | HDP: Any HDP | 4 | 2 | 3 | 9 |
| Torrance, 2007(138) | South Africa | cohort | no | 187 | 1 | 30.4 | SGA | SGA: BW<P10 | 4 | 2 | 3 | 9 |
| Trevisanuto, 2013(139) | Italy | ca-co | yes | 98 | 1 | 27.8 | CA | CA: Histologic | 4 | 2 | 3 | 9 |
| Tsai, 2015(140) | Taiwan | ca-co | yes | 1680 | 21 | 26.0 | SGA | SGA: BW<P10 | 4 | 2 | 3 | 9 |
| Turunen, 2011(141) | Finland | cohort | yes | 36 | 1 | 26.6 | HDP | HDP: PE | 4 | 2 | 3 | 9 |
| Upadhyay, 2015(142) | USA | cohort | no | 212 | 1 | 24.0 | Mortality | SGA: BW<P10 | 4 | 0 | 3 | 7 |
| Ushida, 2020(143) | Japan | cohort | yes | 21005 | 200 | 28.2 | HDP | HDP: Any HDP | 4 | 1 | 3 | 8 |
| Van Vliet, 2012(144) | Netherlands | cohort | yes | 72 | 1 | 29.0 | CA | CA: Histologic | 4 | 2 | 3 | 9 |
| Vinnars, 2015(145) | Sweden | cohort | yes | 167 | 7 | 24.5 | CA | CA: Histologic | 4 | 2 | 3 | 9 |
| Wadhawan, 2007(146) | USA | cohort | no | 9461 | 19 | 26.1 | SGA | SGA: BW<P10 | 4 | 2 | 3 | 9 |
| Watterberg, 1999(147) | USA | cohort | yes | 40 | 1 | 25.4 | CA | CA: Histologic | 4 | 1 | 3 | 8 |
| Westby Wold, 2009(148) | Norway | cohort | no | 365 | 15 | 30.2 | SGA | SGA: BW<P5 | 4 | 1 | 3 | 8 |
| Wilmink, 2019(149) | Netherlands | cohort | no | 273 | 1 | 29.3 | HDP | HDP: PE | 4 | 2 | 3 | 9 |
| Wirbelauer, 2010(150) | Germany | cohort | no | 49 | 1 | 28.5 | SGA | SGA: BW<P10 | 4 | 0 | 3 | 7 |
| Wocadlo, 1994(151) | Australia | cohort | no | 438 | 1 | N | Mortality | SGA: BW<P10 | 4 | 2 | 3 | 9 |
| Woo Lim, 2015(152) | South Korea | cohort | no | 1649 | 60 | N | Mortality | SGA: BW<P10 | 4 | 2 | 3 | 9 |
| Xie, 2015(153) | China | cohort | no | 371 | 1 | 31.1 | CA | CA: Histologic | 4 | 2 | 3 | 9 |
| Yu, 2011(154) | Korea | cohort | yes | 415 | 1 | 28.9 | SGA | SGA: BW<P3 | 4 | 2 | 3 | 9 |
| Zanardo, 2010(155) | Italy | cohort | no | 287 | 1 | N | CA | CA: Histologic | 4 | 2 | 3 | 9 |
| Zaw, 2003(156) | Canada | cohort | no | 999 | 1 | 24.8 | SGA | SGA: BW<P10 | 4 | 2 | 3 | 9 |

GA: gestational age; USA: United States of America; HDP: gestational hypertensive disorders, PE: preeclampsia; HELLP: syndrome in pregnancy characterized by hemolysis, elevated liver enzymes and low platelet count; CA: chorioamnionitis; Ca-co: case-control study; SGA: small for gestational age as defined by birth weight percentile; BW: birth weight; P3: 3rd percentile; P5: 5th percentile P10: 10th percentile; IUGR: intra-uterine growth restriction defined on the basis of fetal growth assessment.

## MOOSE Checklist for Meta-analyses of Observational Studies

| Item No | Recommendation | Reported on Page No |
| --- | --- | --- |
| Reporting of background should include |  |  |
| 1 | Problem definition | 3-4 |
| 2 | Hypothesis statement | - |
| 3 | Description of study outcome(s) | 4-5 |
| 4 | Type of exposure or intervention used | 4 |
| 5 | Type of study designs used | 4, Suppl (page 3-4) |
| 6 | Study population | 4 |
| Reporting of search strategy should include |  |  |
| 7 | Qualifications of searchers (eg, librarians and investigators) | Title page, Suppl (page 3) |
| 8 | Search strategy, including time period included in the synthesis and key words | 4, Suppl (pages 1-3) |
| 9 | Effort to include all available studies, including contact with authors | 4, Suppl (page 4) |
| 10 | Databases and registries searched | 4 |
| 11 | Search software used, name and version, including special features used (eg, explosion) | Suppl (page3) |
| 12 | Use of hand searching (eg, reference lists of obtained articles) | Suppl (page 3) |
| 13 | List of citations located and those excluded, including justification | Suppl Table 1, Suppl Fig 1 |
| 14 | Method of addressing articles published in languages other than English | Suppl (page3) |
| 15 | Method of handling abstracts and unpublished studies | Suppl (page3) |
| 16 | Description of any contact with authors | Suppl (page 4) |
| Reporting of methods should include |  |  |
| 17 | Description of relevance or appropriateness of studies assembled for assessing the hypothesis to be tested | 4 |
| 18 | Rationale for the selection and coding of data (eg, sound clinical principles or convenience) | 4 |
| 19 | Documentation of how data were classified and coded (eg, multiple raters, blinding and interrater reliability) | 4 |
| 20 | Assessment of confounding (eg, comparability of cases and controls in studies where appropriate) | 4 |
| 21 | Assessment of study quality, including blinding of quality assessors, stratification or regression on possible predictors of study results | 4 |
| 22 | Assessment of heterogeneity | 4-5 |
| 23 | Description of statistical methods (eg, complete description of fixed or random effects models, justification of whether the chosen models account for predictors of study results, dose-response models, or cumulative meta-analysis) in sufficient detail to be replicated | 4-5 Suppl (pages 3-4) |
| 24 | Provision of appropriate tables and graphics | Table 1 Suppl tables 1-6 Figs 1-3 Suppl Figs 1-8 |

| Reporting of results should include |  |  |
| --- | --- | --- |
| 25 | Graphic summarizing individual study estimates and overall estimate | Figs 1-2<br>Suppl Figs 2-7 |
| 26 | Table giving descriptive information for each study included | Suppl Table 1 |
| 27 | Results of sensitivity testing (eg, subgroup analysis) | Fig 1-2, Suppl Figs 2-7<br>Suppl Tables 2, 4 and 5 |
| 28 | Indication of statistical uncertainty of findings | Figs 1-2<br>Suppl Figs 2-7 |

| Item No | Recommendation | Reported on Page No |
| --- | --- | --- |
| Reporting of discussion should include |  |  |
| 29 | Quantitative assessment of bias (eg, publication bias) | 8, Suppl Fig 8 |
| 30 | Justification for exclusion (eg, exclusion of non-English language citations) | Suppl (page 3) |
| 31 | Assessment of quality of included studies | Suppl Table 1 |
| Reporting of conclusions should include |  |  |
| 32 | Consideration of alternative explanations for observed results | 7-10 |
| 33 | Generalization of the conclusions (ie, appropriate for the data presented and within the domain of the literature review) | 7-10 |
| 34 | Guidelines for future research | 7-10 |
| 35 | Disclosure of funding source | 11 |

*From:* Stroup DF, Berlin JA, Morton SC, et al, for the Meta-analysis Of Observational Studies in Epidemiology (MOOSE) Group. Meta-analysis of Observational Studies in Epidemiology. A Proposal for Reporting. *JAMA*. 2000;283(15):2008-2012. doi: 10.1001/jama.283.15.2008.

